# Hybrid Health Recommender Systems for Antibiotic Prescription in Sepsis Patients: Statistical Method Development and Evaluation

**DOI:** 10.1101/2025.11.21.25340749

**Authors:** Hannah Marchi, Sophie Schmiegel, Rainer Borgstedt, Sebastian Rehberg, Christiane Fuchs

## Abstract

**Background:** Patient survival in sepsis critically depends on immediate and appropriate empiric antibiotic therapy. Selecting the optimal therapy is challenging in practice due to often incomplete information and time constraints, while minimizing broad-spectrum antibiotic use is essential to prevent resistance development. Clinical decision support systems (CDSS) can assist physicians in deciding on an empiric therapy.

**Methods:** We explore health recommender systems (HRS) as a methodological basis for such CDSS with the aim to predict the therapy response of newly diagnosed sepsis patients to all eligible therapies. Using retrospective clinical and microbiological data from a German university hospital, we introduce and evaluate four HRS of varying complexity, input information and applicability. Each system combines collaborative and demographic filtering in a hybrid manner, employing both memory- and model-based filtering techniques. The HRS make use of both patient similarity (defined by core data and laboratory values) and therapy similarity (defined by pathogen coverage).

**Results:** The best results regarding precision, and F1 score are achieved by the least complex HRS (using memory-based filtering with patient similarity) and the most complex HRS (using model-based filtering and integrating patient core data and laboratory values). However, performance varies significantly across individual therapies. Memory-based methods are particularly suitable for new patients without prior therapy information if data is balanced, whereas model-based approaches perform better when therapy response is already partially available.

**Conclusions:** We present a methodological framework for hybrid HRS that aligns both memory-and model-based approaches with their respective optimal clinical scenarios, while underscoring the critical necessity for high-quality data to handle patient heterogeneity. While application in clinical practice is not yet feasible based on current data, our proposed approaches advance sepsis therapy research and offer a foundation for future studies.

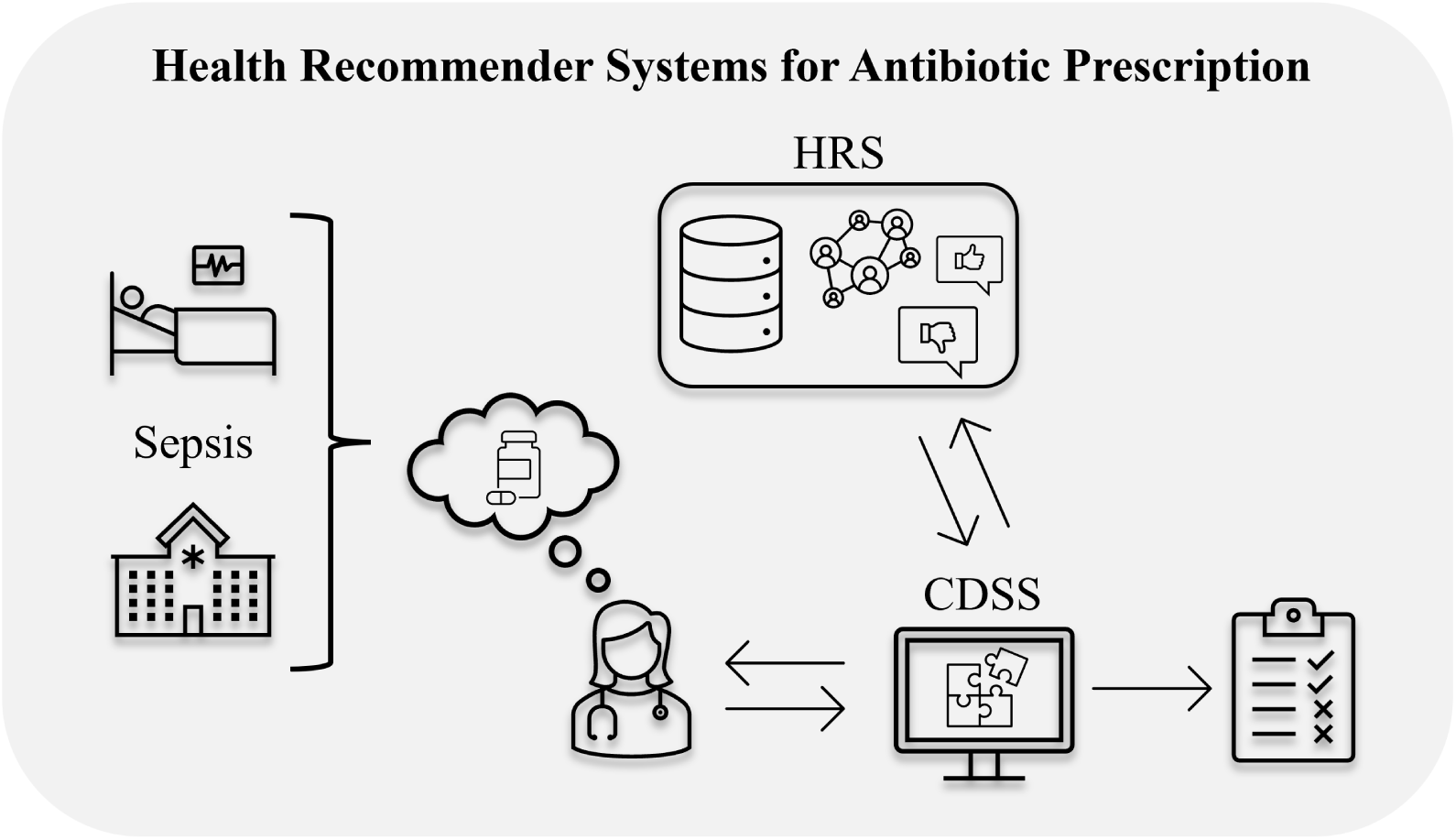

## 1 Background

### 1.1 Introduction

Sepsis is a life-threatening condition. A total of around 49 million cases of sepsis were recorded on a global scale in 2017, resulting in 11 million deaths and accounting for 19.7 % of global mortality (Rudd et al., 2020). Even in case of survival, sepsis patients often suffer from considerable short-and long-term consequences (Vincent, 2025). Antibiotic treatment within the first few hours after diagnosis is crucial for patient survival and forms part of established sepsis treatment guidelines (Evans et al., 2021). However, broad-spectrum antibiotics, especially, should be used judiciously to prevent the development of antibiotic resistance (Rhee et al., 2020). Nevertheless, valid therapy decisions require information such as the present pathogens and other laboratory values, which is only available after several hours or even days. Therefore, due to incomplete information and time constraints, it remains a considerable challenge to rapidly and reliably select an appropriate empiric antibiotic therapy for sepsis patients, even for experienced physicians.

Decision support systems (DSS) are information systems designed to assist users in data analysis and decision-making by providing relevant information, models, and tools through a knowledge-based infrastructure (Hak et al., 2022). Clinical decision support systems (CDSS) represent a subgroup of DSS, introduced in the 1980s, and specifically support patients or healthcare professionals in health-related decisions (Sutton et al., 2020). Data-driven techniques within a CDSS can identify patterns that may be unknown or difficult for humans to process. While a CDSS is designed to enhance decision-making, it is not intended to replace the physician. It aims to improve the overall quality of treatment by providing valuable insights. Nevertheless, clinicians must ensure that the suggested therapies are suitable for the specific patient and that all decisive factors are taken into account. Implementing a CDSS offers a viable solution to assist treating physicians in making timely and informed decisions by recommending the most suitable therapies for each individual patient and could be highly valuable in clinical settings. The development of such a CDSS for sepsis represents the overarching objective of the interprofessional project ‘KINBIOTICS’ (KINBIOTICS, 2025), which the authors contributed to.

The methodological basis of CDSS is formed by integration of statistical models and algorithms, for instance recommender systems (RS; for details about the relationship between (C)DSS and RS see A.3). RS are tools designed to suggest relevant items (such as products, media or services) to users by filtering and predicting user preferences based on various data inputs. These systems have become a standard component of our daily lives and are widely used across different domains. The purpose of employing RS is to enhance the user experience through personalized content and recommendations. The conceptual background of RS differs for applications in healthcare, henceforth referred to as health recommender systems (health RS, HRS). Unlike users who can explore and rate products and adjust their preferences, sepsis patients lack both inherent preferences and the ability to intentionally trial multiple antibiotic therapies. Furthermore, only a limited number of possible therapies are administered to each patient. This results in a small number of therapy entries per patient and consequently sparse data as a basis for the construction of an RS. To address the aforementioned challenge of assisting physicians in identifying the most appropriate antibiotic therapies, we develop several HRS, based on different methodological concepts, for the application in the context of sepsis. We use retrospective microbiology data, core data, vital signs and blood values from previous patients to train the HRS and estimate therapy response for each eligible therapy for patients newly diagnosed with sepsis. These HRS can finally be incorporated into a CDSS for sepsis management to support treatment decision.

### 1.2 Medical background and clinical routines

Since 2016, sepsis has been defined by the ‘Sepsis-3 definitions’ as ‘life-threatening organ dysfunction caused by a dysregulated host response to infection’ (Singer et al., 2016). This condition arises when organisms such as bacteria, viruses, fungi, or parasites enter the bloodstream, prompting the immune system to trigger an immune reaction that leads to systemic inflammation. This widespread inflammation can subsequently result in organ failure and even death. The respiratory tract, particularly in the form of pneumonia, is the most common site of initial infection, followed by unspecified sites, the urinary tract, the abdomen, and the skin (Mayr et al., 2014). There is no doubt that accurately determining the incidence and mortality rates is challenging, and sepsis remains the leading cause of death resulting from infection (Singer et al., 2016).

The international guidelines for management of sepsis and septic shock (Evans et al., 2021) provide detailed guidance for clinicians to treat sepsis patients. Besides other precautions, they recommend to immediately start administration of antibiotic therapy, ideally within one hour of recognition (for patients with suspected septic shock or confirmed sepsis) and within three hours (in case of possible sepsis). In order to select an appropriate antibiotic, a number of aspects have to be considered, including patient history, clinical examination and available blood values and vital signs. Moreover, severity of illness, local resistance prevalence and patient risk factors need to be taken into account. Evans et al. (2021) recommend to use an empiric broad-spectrum therapy with one or more antimicrobials to cover all likely pathogens.

A crucial step for therapy selection is to further culture a patient sample such as blood, urine, or sputum in a laboratory to isolate the bacterial pathogens, and test for resistance towards a panel of antibiotics. This procedure is called antimicrobial susceptibility testing (AST) and reveals both the type of pathogens present and their sensitivity to selected antibiotics. The pathogens are classified as resistant (R), intermediate (I) or susceptible (S) to each antibiotic tested. Once causative pathogens and susceptibilities are known from AST, the broad-spectrum therapy should be de-escalated or changed to more specific antibiotics (Evans et al., 2021). However, AST is cost-intensive and time-consuming. By using current AST techniques, results are available only after 24 to 48 hours, sometimes even up to 72 hours (Gajic et al., 2022). This implies that important information regarding existing pathogens and resistances to certain antibiotics is not yet accessible at the time when an empirical antibiotic must be chosen: this choice relies on information available within the initial hours following diagnosis, such as the patient’s medical history, core data, vital signs, laboratory values, the suspected infection focus, and the physician’s clinical experience. Schmiegel et al. (2025) further explore specific requirements for therapy decisions in sepsis patients, as identified through structured interviews with practicing clinicians.

In the context of sepsis, Rhee et al. (2020) found that both inadequate and excessive use of broad empiric antibiotics are linked to increased mortality rates. Generally, factors such as the overuse and inappropriate prescribing of antibiotics significantly contribute to the rapid development of resistance (Ventola, 2015). According to the findings of Murray et al. (2022), an estimated 4.95 million deaths were associated with bacterial antimicrobial resistance (AMR) in 2019, from which 1.27 million deaths were directly attributable to bacterial AMR. Consequently, to reduce the development of resistances, a critical objective in prescribing antibiotic therapy for sepsis patients is the careful selection and use of targeted antibiotics whenever feasible. However, incorporating information about resistance patterns poses an additional challenge for physicians within the treatment decision-making process.

### 1.3 Related work and research question

To advance the diagnosis, treatment and prevention of sepsis, intense research is being conducted by various subject domains such as immunology, microbiology, medicine and biotechnology. From the field of data science, this includes the analysis of health datasets to gain insights into risk factors, early diagnosis or improved treatment, and thereby revealing unknown patterns that can contribute to the management of sepsis patients. Important contributions, among others, address early prediction of sepsis (Islam et al., 2023, Moor et al., 2021), applications for early detection, real-time monitoring and management of sepsis using methods of machine learning (ML) and artificial intelligence (AI) (Li et al., 2024, Yang et al., 2023), and the prediction of mortality risks (Dong et al., 2024) and antibiotic resistance (Cohen et al., 2025, Ferrari et al., 2024, Vouriot et al., 2024) in sepsis patients using integrated network models and (interpretable) ML.

Regarding sepsis treatment, recent research is being conducted to optimize drug dosages using deep neural networks (Baek, 2023). Liu et al. (2023) aim to identify the effective treatment timing, and Weinberger et al. (2020) analyze the relationship between time-to-antibiotics and mortality for patients with suspected sepsis. Choi et al. (2024) contribute to improving sepsis treatment through deep learning that is able to extract an optimal treatment policy (including dosage and vasopressor treatment) from treatment records, focusing on the patients’ estimated survival rates.

Recent developments of CDSS for medical treatment (in a broader context than sepsis or antibiotics) include Verboven et al. (2022), Cho et al. (2024), Acosta-García et al. (2025) and Ghosh et al. (2025). Regarding CDSS for antibiotic prescription, the TREAT project employs causal probabilistic networks for treating bacterial infections while reducing antibiotic costs and the use of broad-spectrum antibiotic treatment (Leibovici et al., 2007, Paul et al., 2006) and evaluate the system on patients with sepsis (Paul et al., 2007). However, the primary focus of this study is the prediction of the probability for pathogens to be present, with the subsequent recommendation of therapies based on these predictions. Unlike in our case, the study exclusively considers patients in whom the focus of infection is evident. Aside from that project, Delory et al. (2020) and Schaut et al. (2022) present guideline-based rather than data-driven CDSS for antibiotic prescription. Parra-Rodriguez and Guillamet (2022) provide a review on questions about antibiotic decision-making in intensive care units (ICU) — whether to start antibiotic therapy, which spectrum and possible de-escalation — and the role of ML and AI in CDSS.

This plethora of research demonstrates the relevance and complexity of choosing the optimal therapy, and that researchers are approaching the solution based on specific initial conditions. To the best of our knowledge, there is no implementation of a CDSS in clinical practice which is capable to reliably recommend an empiric antibiotic at the time of sepsis diagnosis for patients with unconfirmed infection focus. Such a CDSS may consist of various components, including recommendations for appropriate therapies, information on potential side effects, consideration of individual patient needs, monitoring tools, and alarm systems. With our work, we strive for contributing to the first component: the recommendation of suitable therapies based on their predicted therapy response. To this end, we build upon our previous study (Schmiegel et al., 2025) in which we aimed to predict the prescribed empiric antibiotics for sepsis patients using retrospective data. While that work is particularly useful for the exchange of experience between practicing clinicians and the evaluation of prescriptions, it is limited for the intended use here in the context of a CDSS: the documented treatment choices reflect the decisions made based on limited information and under time pressure. We posit that these do not always align with the optimal therapy that could have been selected under more beneficial conditions. Therefore, we extend the scope of our research from merely predicting the administered therapy to predicting the most appropriate empiric antibiotic therapies for patients newly diagnosed with sepsis. With the current work we address the research question of developing a capable method that meets the specific requirements for the medical context. The resulting approach is intended to serve as a key component within a broader CDSS for sepsis management.

The remaining article is structured as follows: Section 2 details the methods employed for our analysis. We describe the data used in this study in Section 2.1. In Section 2.2, we define the requirements and assumptions to address the research question and motivate the application of RS. Subsequently, in Section 2.3, we give an overview over the general concept and common filtering techniques, that are methods used to select from a large set of data only those elements that meet certain criteria. We then transition to HRS for sepsis treatment in Section 2.4, where we develop HRS approaches for application to the considered context. This includes the consideration of how to evaluate RS results. In Section 3, we report the results of our analysis, in which we applied the presented methods to a patient-derived clinical dataset. Finally, we discuss our findings in Section 4, and we summarize and conclude our work in Section 5.

## 2 Methods

### 2.1 Data

The data used in this study was obtained from patients who were admitted to the adult ICU of the Protestant Hospital of the Bethel Foundation, a university hospital in Bielefeld, Germany, between 2012 and 2023. All patients were diagnosed with either sepsis or septic shock. The patient data comprises time-constant core data, longitudinal data for blood values and vital signs measured over the period of hospitalization and microbiology data, including categorical results of the AST. A detailed description of the original dataset and relevant preprocessing steps is provided in our previous work (Schmiegel et al., 2025), where we identified and demonstrated essential data preprocessing steps for developing a CDSS for therapy decisions in sepsis patients. In order to address the research question of this work, additionally to that preprocessing, we undertook further steps delineated in Section A.1.1, including the reduction and categorization of specific variables and the imputation of missing values. In order to concentrate on the early time point at sepsis diagnosis for prediction purposes, we reduced the longitudinal data such that there is only one value for each measurement. To this end, we took the earliest measurement within the first hour after diagnosis, if available, and otherwise the most recent measurement within 24 hours before diagnosis. Microbiology data includes AST results, i. e. infection ID, the sample substance, the present pathogen and the resistance status (S/I/R) for selected antibiotics per patient. During preprocessing, we adjust and filter the AST data and build combined therapies according to the most frequently prescribed combinations. We further reduce these therapies according to rules defined in Section A.1.1.

The variable ‘infection ID’ represents a combination of patients and infections since some patients are represented multiple times: they were either admitted to the hospital more than once, or they had multiple infections during one hospital stay. In the following, we use the term ‘patients’ to refer to the combination of patient and infection, especially when indicating sample sizes (Schmiegel et al., 2025). The final dataset for our analysis contains 703 patients. Table 1 shows all included variables from core data, laboratory values and vital signs. The patients’ age ranges from 18 to 99 years with an average of 67.3 years; the proportion of female patients is 36 %. The amount of missing data per variable lies between 0 % and 31.01 % (mean 9.89 %); Figure A1 illustrates missing measurements for all patients per variable.

**Table 1:**
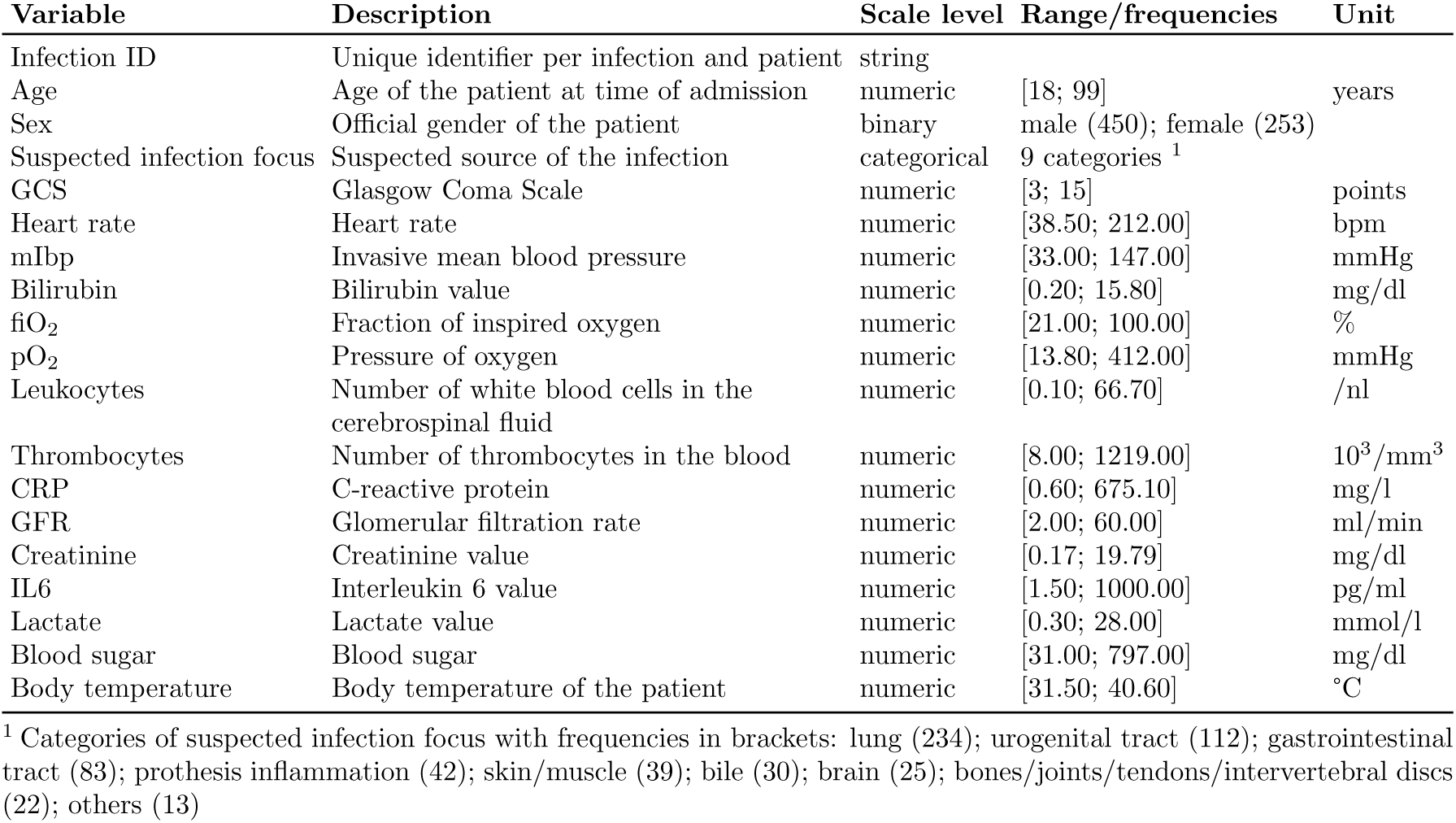
Variables from core data, laboratory values and vital signs contained in the final dataset from n = 703 patients.

Microbiology data is exemplarily shown in its original form in Table 2. Missing values (displayed as ‘NA’) in the resistance status indicate that the antibiotic in question was not tested in the AST for the specified patient. In the majority of cases, there is more than one detected pathogen per patient, and thus there appear multiple rows per infection. For the CDSS, we require one single result per patient per therapy (across all detected pathogens). We achieve this by summarizing the non-missing resistance information for each therapy and per patient as follows: The ultimate outcome is considered resistant if there is at least one pathogen which is resistant to the given therapy; otherwise, it is labeled susceptible. The microbiology data contains resistance information from all 703 patients for 22 single antibiotic therapies. All therapies and according frequencies of resistant and susceptible AST results are listed in Table A1; a graphical representation is shown in Figure A2. Having summarized the AST results to one entry per infection, there are 39.3 % combinations of patient and therapy without any AST result, 41.1 % with result ‘susceptible’ and 19.6 % with result ‘resistant’. The therapy that was tested in most patients (622/703) is Cotrimoxazol, with ‘Penicillin G’ tested least frequently (202/703). The therapy with the highest proportion of susceptible results is Oxacillin (92.8 %), and the one with highest proportion of resistant results is Ampicillin with 74.1 %. We observe a considerable imbalance in the distribution of resistance categories across therapies, primarily with a majority of class ‘susceptible’ and a minority of class ‘resistant’. In certain instances, such as with Ampicillin or Piperacillin, this imbalance is inverted.

**Table 2:**
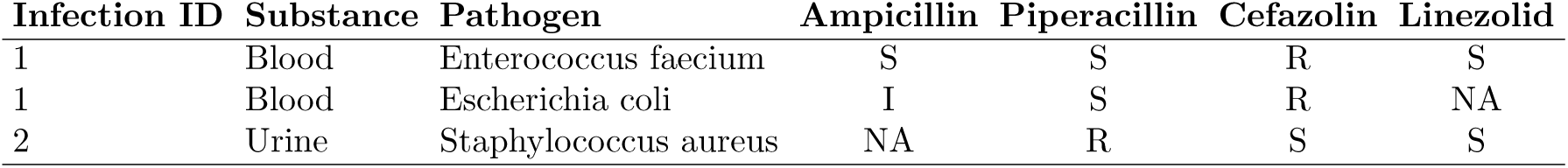
Example data from AST. For each patient, several rows may appear, representing testing for different pathogens. Columns headed with names of antibiotics represent the resistance status of the pathogen to the specific therapies, defined as R: resistant, I: intermediate, S: susceptible, NA: not tested.

| Infection ID | Substance | Pathogen | Ampicillin | Piperacillin | Cefazolin | Linezolid |
| --- | --- | --- | --- | --- | --- | --- |
| 1 | Blood | Enterococcus faecium | S | S | R | S |
| 1 | Blood | Escherichia coli | I | S | R | NA |
| 2 | Urine | Staphylococcus aureus | NA | R | S | S |

Further model-specific preprocessing steps, including imputation of missing values and handling of class imbalance, are described in Section A.2.1.

### 2.2 Statistical Method

At the core of a CDSS lies a statistical method for incorporating and analyzing the available data. The unique characteristics of this data, along with the medical context, impose specific requirements for the method that must be addressed, including data properties, modeling assumptions and practical requirements.

The context of our research question requires a method that is appropriate to suggest antibiotic therapies for patients newly diagnosed with sepsis for whom no prior therapy response information is available. Ideally, it provides a set of therapies rather than a single choice, supporting physicians to select the most appropriate option against the background of their expertise. This allows for the consideration of factors such as side effects, intolerances/allergies and comorbidities.

In order to fulfill these requirements, we implement a procedure that estimates the response to each available therapy for each patient. The challenge lies in the fact that, for new patients, only core data and, at best case, vital signs and blood values can be assumed to be available; valuable information about present pathogens and resistances, in contrast, will not be provided at the time of decision-making.

Statistical estimation of therapy response will be learned on retrospective health records of other sepsis patients. To that end, we rely on the central assumption that patients with similar health records will respond similarly to the same therapy. Vice versa, we assume that therapies with a similar pathogen coverage have a similar effect on the same patient. Furthermore, we assume, as supported by Singer et al. (2016), that the response to therapy is not exclusively influenced by the present pathogens, but also by the patients’ individual health profiles.

With regard to the characteristics of the data, several points require further attention: from the plethora of antibiotic therapies, each patient can only receive a small number of these during one hospital stay (in our dataset, there are up to eleven different therapies per patient, with a mean and median of two). A ‘trial and error’ approach is inconceivable for patients with sepsis, as this would endanger their survival. As a result, there are many missing values for non-administered therapies, and even for the administered ones, it remains unknown whether an alternative therapy would have been equally or more effective than the one received. Furthermore, the frequency with which particular therapies are prescribed varies substantially, leading to a considerable imbalance in the number of patients receiving them.

Regarding the translation into clinical practice, physicians from our KINBIOTICS consortium stressed the importance of understanding the systems’ suggestions to comprehend the decision-making process, which also contributes to the system’s trustworthiness (Schmiegel et al., 2025). Our method selection is thus guided by finding a compromise between prediction performance and interpretability for the specific application of treatment for sepsis patients.

Taking all aspects together, we are approaching the solution to the problem of suggesting suitable antibiotic therapies through the development of an HRS. The concept of RS has been developed, refined, and applied in numerous contexts, particularly outside the healthcare domain. Section 2.3 provides a general introduction in order to highlight strengths, weaknesses, and limitations. Building on these foundations, Section 2.4 develops specific approaches tailored to the context of this work.

### 2.3 Recommender systems

Recommender systems filter datasets for relevant information in order to generate personalized recommendations for items (such as products or services) to subjects (such as users) based on previously recorded data. Everyday examples of RS are web shops like Amazon (Amazon, 2025) or audio streaming platforms like Spotify (Spotify, 2025), where (new) products or songs are suggested to customers based on their user profiles, interests and previously purchased products/songs. The underlying assumption of RS is the presence of substantial dependencies between subject- and item-related activity. That is to say, within certain groups, people are assumed to have similar preferences. At the same time, people are assumed to generally like items that are similar to those they have previously purchased or consumed (Aggarwal, 2016). The system leverages existing information from subjects on items, such as ratings or behavior, to make appropriate suggestions and filter relevant information for individual users (Ricci et al., 2022). By providing personalized suggestions to users, RS help to overcome information overload and to better understand user preferences, resulting in a more engaging user experience and higher satisfaction (Isinkaye et al., 2015).

Different contexts require different data and background information to be incorporated into the RS. However, there is a common structure for all types of RS, including three essential elements: **users**, **items** and an evaluation measure, in the following called **rating** (Ricci et al., 2022):

- **Users** are individuals that are consuming any kind of product or information with different objectives such as customers of shopping platforms, users of streaming services or visitors of restaurants. Depending on the type of RS, user characteristics are incorporated in the decision process to provide personalized recommendations. In addition, user relationships are taken into account, for instance to find groups of similar users.
- **Items** are the elements that are evaluated and recommended in an RS. Examples include products (e. g. books, cars or food), entertainment (e. g. music or movies), content (news, web articles or e-books) or services (restaurants, travel, accommodation, service providers etc.). Items can have characteristics of varying complexity such as genre, year of publication and number of pages in the case of books or the price and terms and conditions of insurance policies (Ricci et al., 2022).
- **Ratings** are a user’s evaluation of items. These can be direct measures, such as explicit preferences, opinions and ratings, or indirect or implicit measures, such as observed behavior. Hybrid measures combine implicit and explicit feedback (Isinkaye et al., 2015). Ratings can be numeric (e. g. 1-5 stars rating scale), ordinal (using ordered categorical values, e. g. good’, ‘neutral’, ‘bad’), binary (e. g. ‘agree/disagree’) or unary (possibility to like but not to dislike) (Ricci et al., 2022).

These three elements form the foundation of a so-called user-item matrix or rating matrix ***R*** ∈ R*^n^*^×^*^m^*, where *n* is the number of users and *m* is the number of items. The vector ***p*** = (*p*_1_, · · ·, *p_n_*) contains the users and ***t*** = (*t*_1_, · · ·, *t_m_*) is the vector of all available items. Each element of ***R*** is filled with the rating *r_i,j_* from user *p_i_* on item *t_j_* (with *i* = 1, · · ·, *n* and *j* = 1, · · ·, *m*) if available, or left empty otherwise (see Figure 1a). This matrix ***R*** is used as data basis in many RS. In particular, RS aim to identify items with high(est) rankings to individually recommend them to users. They start from the point that ***R*** is incomplete, containing for each user only ratings for some of the items (usually different ones for different users), indicating the presence of unexplored user-item experiences. The RS fills the missing ratings of ***R*** with estimated ratings *r̂_i,j_* by leveraging the available information (see Figure 1b).

**Figure 1:**
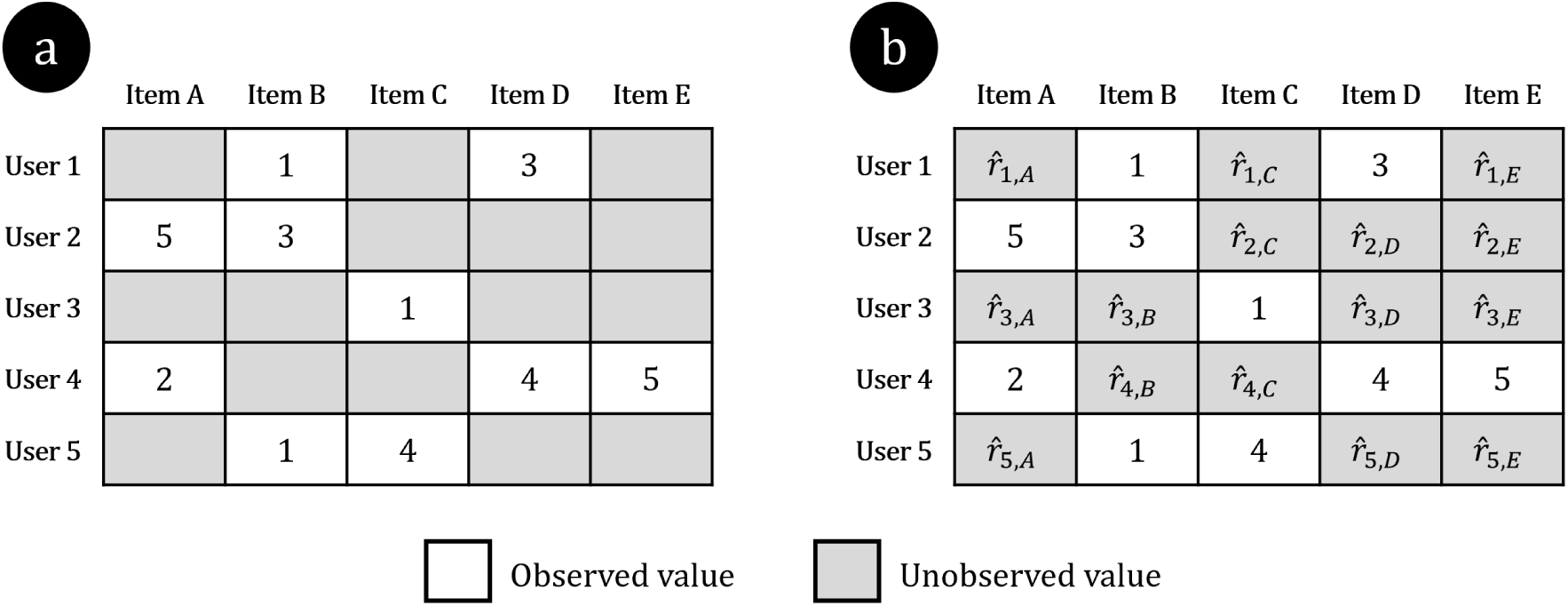
Examples of a user-item matrix ***R*** with interval-based ratings from five users on five items. **a**: Incomplete matrix — if a user has not given a rating on a particular item, there is no entry in the matrix for this user-item combination. **b**: Complete matrix — previously empty cells are filled with estimated ratings, which are then used to recommend to each user those items for which the individually predicted ratings are highest.

According to Aggarwal (2016), distinguish between two general ways how the recommendation problem of an RS can be formulated: the matrix completion problem (MCP) and the top-k recommendation problem (KRP). MCP can be considered a specific instance of data imputation, particularly in the context of a large and sparse underlying data matrix (Aggarwal, 2016). It must, however, be taken into account that missing entries of ***R*** are likely to not be missing at random. In addition, the order of predicted rankings is often more important than the predicted value itself. In line with that, KRP recommends a list of the top *k* items, with *k* specified by the modeler. Rather than predicting exact ratings for items, KRP aims to directly recommend the top items. Once the MCP has been solved, KRP can easily build on it by sorting all rankings per user and selecting the top *k* items. For large *m*, MCP is computationally much more costly.

Different filtering techniques can be applied to construct an RS. In order to illustrate the broad range of possibilities, and to motivate our proceeding, we provide a comprehensive overview of the most frequently used techniques. Further details, including individual strengths and weaknesses, can be found e. g. in Isinkaye et al. (2015). For our analyses, we will eventually use collaborative and demographic filtering, combined into a hybrid approach.

**Content-based filtering (CBF)** focuses on individual users: recommendations are generated based on the user’s own previous ratings. The system suggests items that are similar to those highly rated, purchased or consumed by the same user before. Similarity between items is based on item attributes or features (Ricci et al., 2022). As an example, a user who has previously purchased several regional crime novels with a female protagonist might be suggested further books of the same genre. It is important to note that this approach does not incorporate information from other users when making recommendations (Isinkaye et al., 2015).

As Isinkaye et al. (2015) state, content-based approaches have several advantages: since items are characterized by their attributes, new items can be recommended to users without having been rated by any user beforehand. CBF can quickly adapt to changes in user preferences; the user’s profile does not need to be shared with the system or the modeler, which supports privacy; and it is possible to transparently explain how recommendations were generated. Conversely, a limitation of CBF is that recommendations are contingent on the item’s metadata, and the recommendations for items tend to be similar to items that have already been rated. This prevents innovation.

**Collaborative filtering (CF)** is based on using either the inter-item or inter-user correlation of observed ratings to infer missing ones (Aggarwal, 2016). CF employs either user or item similarity metrics based on relevant common interests and expressed preferences or features (Isinkaye et al., 2015). It makes use of the user-item matrix to suggest items that have not yet been consumed or seen by the user, but have been rated by users in the neighborhood (i. e. a group of users with comparable characteristics). The underlying assumption is that users who share similar interests will prefer similar items. Collaborative techniques are further categorized into two distinct subfields: memory-based and model-based techniques. The main difference between these is the set of users (or items) on which new predictions are based:

- **Memory-based methods**, also known as neighborhood-based methods, are used to compute similarities between users (user-based) or between items (item-based) in order to determine their neighborhood of similar users or items, respectively (Aggarwal, 2016). The existing ratings of these neighbors are aggregated using various statistics (such as a weighted average) to predict values of missing ratings for the user or item of interest (Isinkaye et al., 2015). Memory-based methods are generally considered straightforward to implement and easy to interpret (Aggarwal, 2016).
- **Model-based methods** use information of all available users and items as input for machine learning methods or data mining techniques in order to learn a model for predicting values of missing ratings. Model-based approaches include rule-based models, dimensionality reduction techniques, latent factor models, Bayesian methods or regression (Aggarwal, 2016, Isinkaye et al., 2015).

CF offers several advantages (Isinkaye et al., 2015): it is independent from explicit information about item features, allowing recommendations to be made with no or few information about items. Additionally, CF can provide serendipitous recommendations, i. e. recommendations for novel items that would not have been considered otherwise. However, CF also imply difficulties, such as a reduced system performance caused by the cold-start problem, i. e. the challenge to generate recommendations to new users without prior ratings. Further, CF struggle with data sparsity and item class imbalance. Recommendations tend to be less reliable for items which are rated by only a few users or for users with few similar users. Lastly, CF can become computationally demanding with large datasets as it involves calculating pairwise similarities between users or items. There are further filtering techniques that differ from CBF and CF mainly in the source of information that they use to make recommendations. Three such techniques are presented by Ricci et al. (2022): **Community-based filtering** (also known as social recommender systems) uses the ratings of the user’s friends to infer preferences for items. Conversely, in the case of **demographic filtering**, the recommendations are based on a user’s demographic profile, which may include details such as their origin, native language, gender, and age. **Knowledge-based filtering**, in comparison, use domain knowledge, including a user’s needs and preferences for item features and their utility, to make highly personalized recommendations. Knowledge-based systems are further divided into constraint-based (users explicitly specify their requirements) and case-based (specific scenarios are selected and similar items must be found) techniques.

Each described technique comes with both advantages and restrictions. Combining the techniques in a **hybrid RS** can leverage the benefits of each and mitigate some weaknesses. As outlined by Aggarwal (2016), three designs for hybrid models are distinguished. The ensemble design is applying multiple RS and combining their results into a single output. This combination can be achieved through e. g. weighted averages or sequential ensemble algorithms. Rather than combining the outputs of separate systems, the monolithic design integrates multiple recommendation techniques into a single, unified algorithm by using various data types. Mixed systems, on the other hand, use several separate recommender algorithms, similar to ensembles. However, they present the combination of recommendations from all algorithms as the final result. More details on the principles, benefits, applications and various techniques of RS can be found in the works of Ricci et al. (2022), Aggarwal (2016) and Isinkaye et al. (2015).

An important subfield of RS are HRS. As healthcare becomes increasingly digitized, the amount of available information is growing exponentially. This has led to challenges for patients and practicing clinicians in terms of keeping track of information, filtering relevant data and drawing meaningful conclusions. The complexity of interdependencies present in the available data further exacerbates these difficulties. At the same time, there is a growing interest in personalized medicine and patient treatment. The purpose of HRS is to process different types of health information with the objective of providing personalized recommendations on **items** such as nutrition, treatment options, medication dosage, physical activity, healthcare professionals or health insurances to various **users**, including patients, health professionals and hospitals (Wiesner and Pfeifer, 2014). The information used in the recommendation process encompasses both individual health data, including the patient’s medical history, prior treatments, and laboratory values, as well as more general data, such as disease definitions, prevalence data, information from public databases, and therapy guidelines.

Recent applications or developments of HRS include the assistance in finding relevant information in a medical environment (de Maria et al., 2020), efficient treatment of COVID-19 (Kuanr et al., 2022), treatment of urinary tract infections (Yoon et al., 2022) and treatment of hypertension (Mai et al., 2023). We build upon the work of Gräßer et al. (2017), who present several HRS techniques for therapy decision for patients suffering from the autoimmune skin disease psoriasis and who extend their approach to a pharmaceutical therapy RS (Gräßer et al., 2022). They apply recommender algorithms with demographic-based filtering as well as collaborative filtering, making use of similarity structures. This approach offers a high level of interpretability since recommendations of the system can be transparently explained, and it is relatively simple to implement and to adapt.

### 2.4 Health recommender systems for sepsis treatment

Our work is based on a use case which seeks recommendations for the most appropriate therapies for patients newly diagnosed with sepsis. The users in the HRS are patients, and the recommended items are choices of therapies in the form of single or combined antibiotic treatment. Accordingly, we construct a **patient-therapy matrix** instead of a user-item matrix, where the ratings *r_i,j_* represent the therapy **response** of patient *p_i_*to therapy *t_j_*. As a quantitative measure for the therapy response of administered therapies is not directly recorded in the data, we derive a value by making use of the available microbiology data, which includes resistance information from AST as categorical results (see Section 1.2). This results in a binary measure for therapy response (0 = ‘resistant’ or 1 = ‘susceptible’; see details in Section 2.1). It is important to note that the resistance information per patient is available for several antibiotics, which are, however, not necessarily congruent with the administered antibiotics. Consequently, all analyses presented in this study are based on AST results rather than the actual antibiotics prescribed. Due to the fact that AST is only done for some of all possible antibiotics per patient, the patient-therapy matrix is rather sparse (see data description in Section 2.1 and Figure A2).

In the following, we introduce four different approaches for HRS which we developed for application in case of sepsis. They differ from one another with respect to the choices of (combinations of) filtering (see below; an overview is presented in Figure A3). The HRS incorporate retrospective health record data (i. e. core data, vital signs and blood values) as well as AST results to gain insight into newly diagnosed patients who do not yet have any rating. The aim is to determine the most appropriate therapies for a given patient. We pay particular attention to therapies that have not or rarely been administered to other patients in order to cover the full range of possible treatment options. To this end, we approach the RS problem as an MCP rather than KRP (see Section 2.3). Consequently, estimates are derived for missing ratings (therapy response) from both previous patients and entirely new patients for each possible therapy.

All four HRS employ similarity structures, either among patients or (additionally) among therapies, in order to derive information on non-tested therapies and for newly diagnosed patients. The general procedure for all approaches is as follows:

1. Take the patient-therapy matrix from previous patients as input.
2. Extend the matrix by empty rows to include new patients without any therapy ratings.
3. Fill all missing cells of the extended patient-therapy matrix (procedure to be explained below).
4. For new patients, rank therapies according to their estimated therapy response.
5. Recommend therapies with highest ranking as suitable therapies for the new patients.

The third step of this process is of greatest importance. At this step, the missing values in the patient-therapy matrix are replaced by predicted values. Section 2.3 has outlined the diversity of methodological approaches, their strengths, weaknesses and applicability. In order to take advantage of the broad spectrum of opportunities, the four proposed HRS vary in terms of complexity, the type of input data utilized, and their relative suitability for specific scenarios. Two of these systems are memory-based and two are model-based.

#### 2.4.1 Memory-based HRS

##### Approach Ia: Hybrid memory-based user-based HRS

Our study starts with a comparably straightforward technique: a hybrid memory-based user-based approach that combines collaborative and demographic filtering, in accordance with the approach of Gräßer et al. (2017) and Gräßer et al. (2022). In brief, the HRS first identifies patients whose ratings may be relevant for a particular new patient, and then estimates therapy response from ratings of these patients.

Gräßer et al. (2017) are working with data for patients suffering from the skin disease psoriasis. In that scenario, a patient has several consultations and relevant therapies up to the time of the consultation under consideration. In contrast, in the context of sepsis, only patient data from the initial time point subsequent to diagnosis is available, and no ratings concerning therapies are available for new patients. In this case, the patient-therapy matrix from previous patients and the patient health records of both previous and new patients are used. Gräßer et al. (2017) are calculating the similarity between several consultations of one patient with the objective to give a therapy recommendation. Conversely, we base our recommendations for a new patient on previous ratings of similar users. This approach is founded on the assumption that patients with comparable health records respond similarly to the same therapies. To calculate similarity between consultations, Gräßer et al. (2017) suggest, among others, using Gower’s similarity coefficient (Gower, 1971), which can handle both mixed type variables as well as missing values. In the context of health records, both aspects are of utmost importance. A further benefit of Gower’s similarity coefficient is its capacity to allocate weights to variables, enabling the user to emphasize specific ones. Overall, the coefficient calculates similarity between two objects for each individual variable and combines these individual scores into a weighted average.

We adapt this procedure from Gräßer et al. (2017), calculating similarity between patients instead of consultations. The individual similarity between patients *p_i_*and *p_l_*(where *i, l* = 1, · · ·, *n*) regarding patient variable *v* from the set of variables *V* is denoted by simP^∗^ (*v*). The specific similarity measure depends on the type of variable *v*. Following Gräßer et al. (2017), the Manhattan distance normalized to the attribute range is applied for similarity computation between ordinal, interval, and ratio-scaled attributes, Jaccard similarity coefficients for dichotomous and simple matching for nominal values. An inclusion value *δ_v,i,l_* is defined as 0 if there is a missing value for covariate *v* for at least one of the patients *p_i_*and *p_l_*, and 1 otherwise. Variable weights *w_v_* assign greater or lower importance to individual variables *v* and can therefore be used for content-related weighting or feature selection. (Section A.3 describes how we choose the weights in our study.) Finally, the similarity simP*_i,l_* ∈ [0, 1] between two patients *p_i_* and *p_l_* is the standardized sum of similarities across all variables:

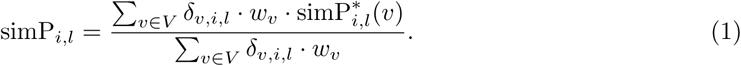

Once the pairwise similarities between patients are calculated, a set *K_i_* of patients similar to patient *p_i_* can be identified. This is done either by specifying the number *ν* of patients with high similarity, or by setting a similarity threshold *ρ* ∈ [0, 1] so that all patients *p_l_* with simP*_i,l_* ≥ *ρ* are considered sufficiently similar. In the following, the threshold is assumed fixed and suppressed in the notation. *K_i,j_* ⊂ *K_i_* defines the set of patients similar to patient *p_i_*who have an existing rating for therapy *t_j_*(with *j* = 1, · · ·, *m*).

For both previous and new patients, missing therapy response is estimated based on ratings from similar patients: the rating *r_i,j_* of patient *p_i_* on therapy *t_j_* is estimated as weighted and normalized average of the (observed) ratings *r_k,j_* from the similar patients *p_k_*∈ *K_i,j_*:

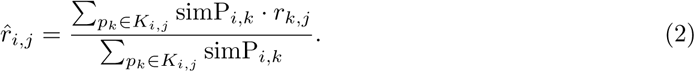

In this manner, all missing entries of the patient-therapy matrix can be filled with predicted ratings. Figure 2 illustrates the procedure of the hybrid memory-based user-based HRS (Approach Ia) using example data.

**Figure 2:**
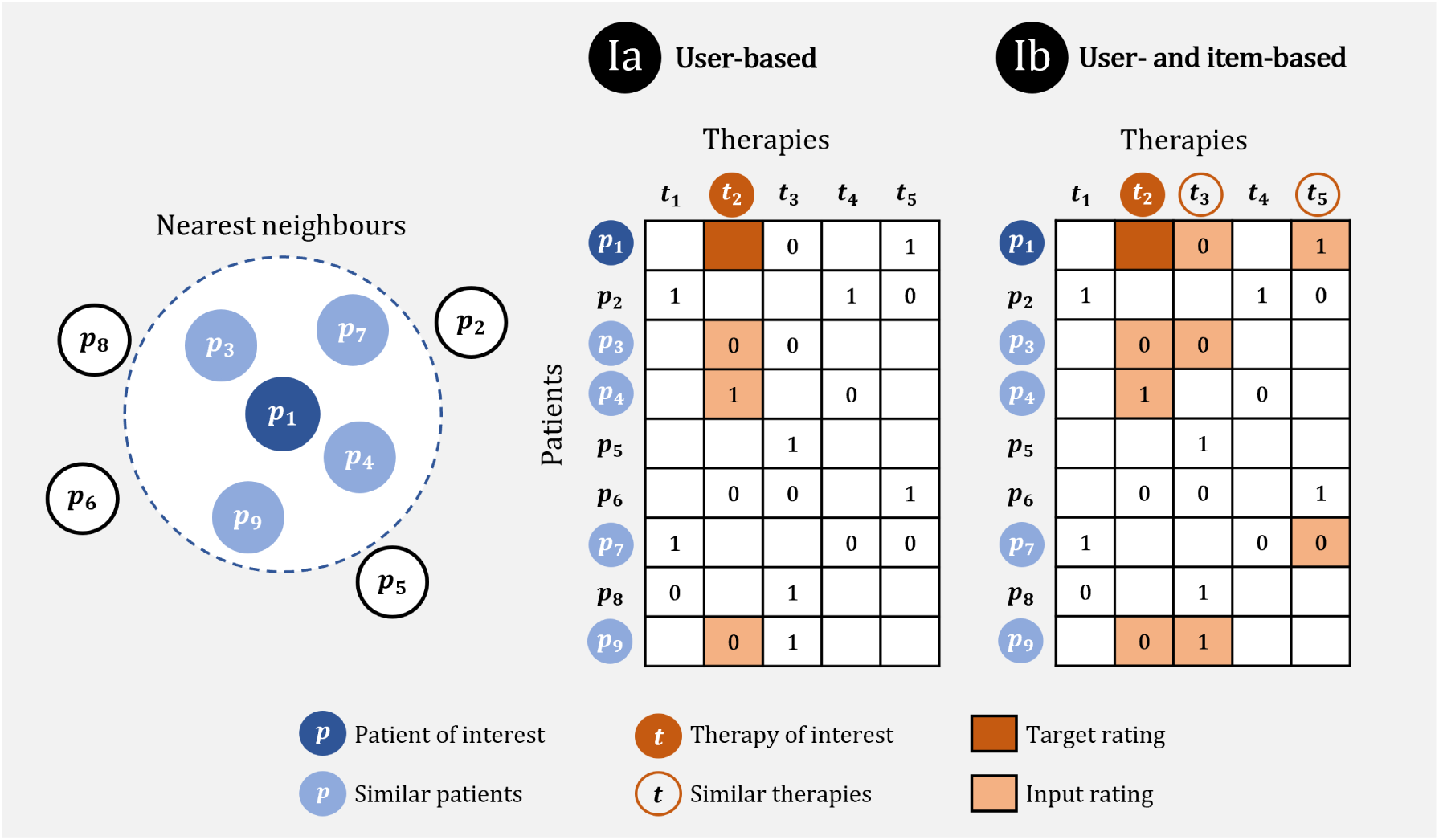
Hybrid memory-based HRS. graphical representation (partially inspired by Gräßer et al. (2022)) of matrix completion in the two hybrid memory-based filtering techniques. In this example, the aim is to predict the therapy response of patient *p*_1_ to therapy *t*_2_. For both HRS, a set of similar patients to *p*_1_ is found. Three out of these patients have an observed rating for therapy *t*_2_. In **Approach Ia** (user-based), the weighted average of these ratings is calculated using Equation (2) to obtain an estimated rating for patient *p*_1_. **Approach Ib** (user- and item-based) additionally considers therapies which are similar to therapy *t*_2_, and the rating for *p*_1_ is estimated by Equation (6). Observed ratings are defined as 0 = ‘resistant’ and 1 = ‘susceptible’.

##### Approach Ib: Hybrid memory-based user- and item-based HRS

The second memory-based approach exploits similarities between patients in the same manner as the first approach (user-based). In addition, it considers that patients will respond similarly to similar therapies (item-based). Therefore, we additionally incorporate ratings from similar patients of similar therapies, thus extending the input values for a specific prediction.

We quantify similarity between therapies using pathogen coverage information: antibiotics differ in their effectiveness in treating specific sets of known pathogens. There are several sources that provide guidelines or coverage information for antibiotic therapy in relation to pathogens. The AMBOSS antibiotic mosaic (AMBOSS GmbH, 2024) shows pathogen coverage for treatment of severe infections, which comprises three coverage categories. We adopt these categories and define the following values for the coverage *c_j_*(*e*) of therapy *t_j_* regarding pathogen *e* from an overall set *E*:

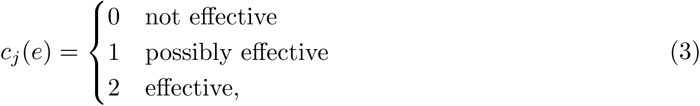

where, based on the AMBOSS antibiotic mosaic, ‘possibly effective’ means that the therapy is in principle effective but with questionable clinical effectiveness or the occurrence of resistance at varying frequencies. The term ‘not effective’ is used for a therapy that is either not effective, not sufficiently investigated or not recommended, while ‘effective’ indicates good effectiveness against the pathogen in consideration. With information on all therapies and pathogens, it is possible to construct a pathogen-therapy matrix (see Section 3). To quantify the similarity simT^∗^ (*e*) between two therapies *t_j_* and *t_h_*(with *j, h* = 1, · · ·, *m*) regarding one specific pathogen *e* ∈ *E*, we propose a variation of the simple matching measure (Šulc and Řezanková, 2019) such that the following assumptions hold: equal coverage (coverage matches) leads to higher therapy similarity; mismatches between coverage category 1 (possibly effective) and 2 (effective) result in a higher therapy similarity than mismatches between categories including 0 (not effective). Specifically, we define

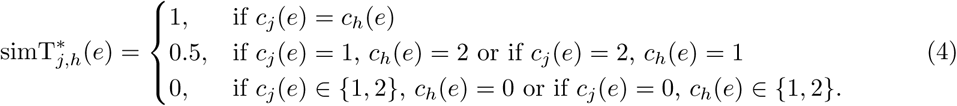

To compare therapies regarding their overall pathogen coverage, we follow suggestions for similarity measures for nominal data from Šulc and Řezanková (2019) and define the total similarity simT*_j,h_* ∈ [0, 1] between two therapies *t_j_* and *t_h_* as the arithmetic mean over all pathogens, which is calculated as:

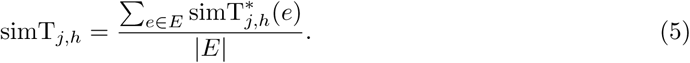

In Approach Ib, the similarity between therapies is included as additional weight in Equation (2). To this end, we set a therapy similarity threshold *τ* ∈ [0, 1] and determine a set *T_j_* of therapies *t_h_* that are similar to *t_j_*, i. e. simT*_j,h_* ≥ *τ*. As for the patient similarity threshold *ρ* before, we suppress the therapy similarity threshold *τ* to not overcomplicate notation. For fixed *ρ* and *τ*, the rating *r_i,j_* of patient *p_i_* for therapy *t_j_* is then estimated as a weighted and normalized average of ratings from patients similar to *p_i_* who were administered a therapy similar to *t_j_*(including *t_j_* itself):

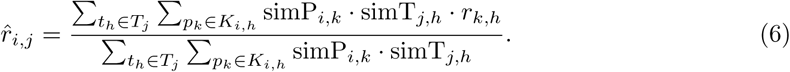

A graphical representation of the procedure is provided in Figure 2.

Overall, it is noteworthy that both memory-based approaches exclusively incorporate observed ratings as input values for estimating unobserved ratings. Details about how patient similarity, therapy similarity and the patient-therapy matrix for Approach Ia and Approach Ib are obtained with our data are described in Section A.3.

#### 2.4.2 Model-based HRS

The structure of the model-based HRS differs from the memory-based approaches mainly in the choice of input ratings and the way of predicting ratings. The first step in the matrix completion process is to fill the empty cells of the patient-therapy matrix with initial values between zero and one. These values may correspond to either row means, column means, or the result of any collaborative filtering technique (Aggarwal, 2016). In subsequent steps, the predictions for ratings are iteratively updated to achieve more reliable values. To that end, we go through all columns of the patient-therapy matrix one after the other and do the following:

1. Consider the column (i. e. the ratings for the respective therapy) as target variable *y* and all other columns as input variables *x*_1_, *…, x_m_*_−1_ for a prediction model M. Split the patient-therapy matrix such that all rows with originally observed (not imputed!) ratings for the target variable form the training data; for rows with originally missing (now imputed) entries in the target variable, a prediction will be carried out. The input variables contain both imputed and actually observed values, both for training and prediction.
2. Update the originally unobserved values of the target variable with predicted values from model M. There are two ways of including updated values for unobserved ratings (both applied in our study):

a. *Immediate*: The predictions for a target value are entered directly into the matrix after the model for one target variable is estimated, and the predicted values are immediately included as input values for the model of the subsequent target variable.
b. *Collective*: The models for all target variables are estimated based on ratings from the previous iteration. Once all the predictions have been made, the patient-therapy matrix is updated and filled with all new predictions for this iteration collectively.

This procedure is repeated until convergence is reached, i. e. until the predictions for missing entries are hardly changing any more. Figure 3 graphically demonstrates the general procedure of matrix completion in model-based HRS, including both the prediction and the iterative step.

**Figure 3:**
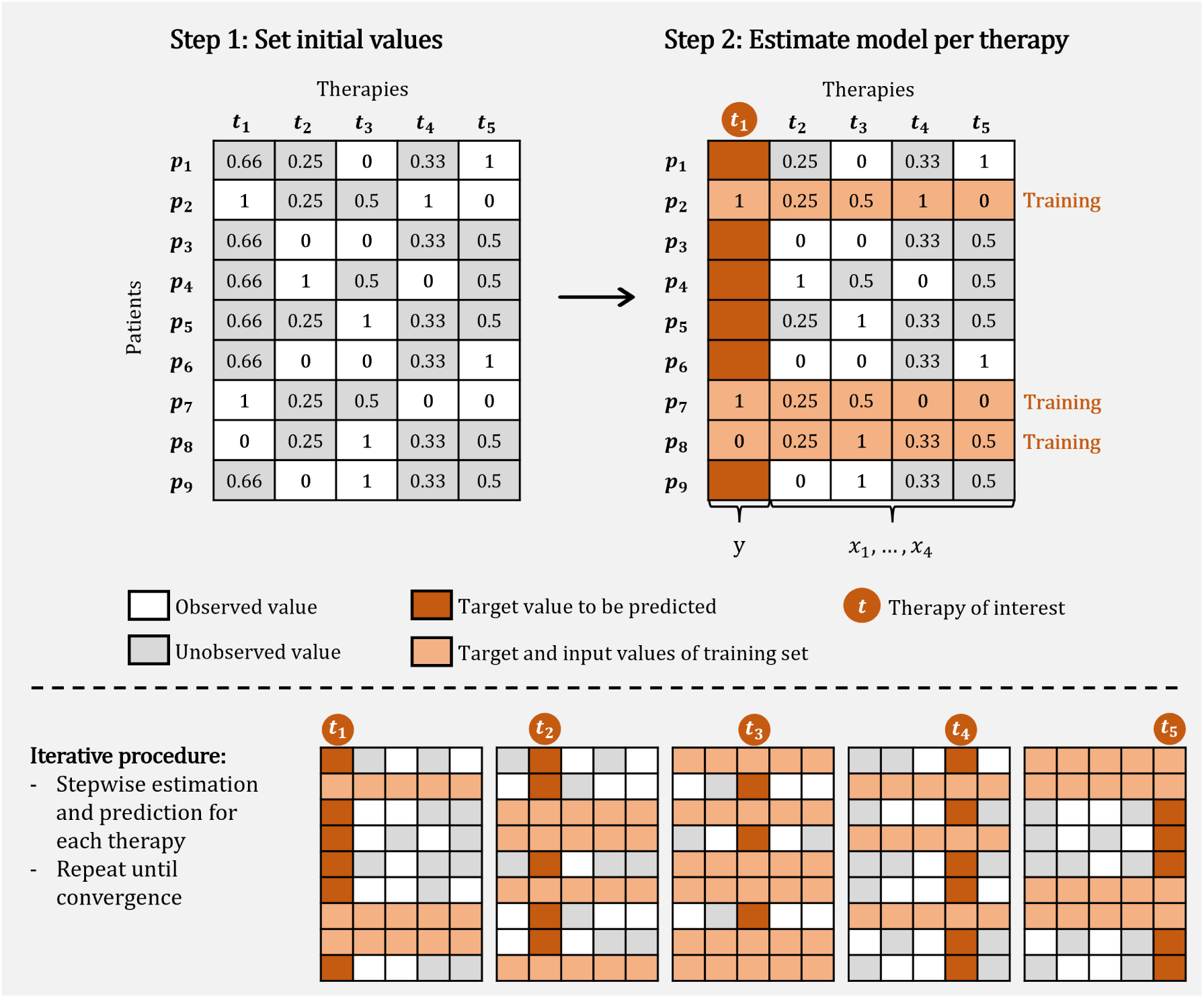
Hybrid model-based HRS. graphical representation of matrix completion procedure in the model-based filtering technique. Initial values (here: column means) are entered into the missing cells of the patient-therapy matrix (observed ratings are defined as 0 = ‘resistant’ and 1 = ‘susceptible’). Subsequently, an iterative procedure is initiated, whereby each therapy is designated as the target variable *y* in a prediction model, with all remaining therapies serving as input variables *x*_1_, *…, x*_4_. For each target variable, the rows with observed ratings are selected as training data, while the rows with missing ratings are assigned for prediction. This iterative procedure is repeated until convergence is achieved.

##### Approach IIa: Hybrid model-based HRS

We combine a model-based HRS with the memory-based approaches from Section 2.4.1 to hybrid filtering. Since the goal is to also predict ratings for patients without any information on therapy response, using mean values per column (i. e. per therapy) as initial values would not be reasonable: in such a scenario, the prediction outputs for all patients would be identical for that therapy. Instead, cells with missing ratings in the patient-therapy matrix are filled with initial values obtained either by Approach Ia or Approach Ib.

In our study, we employ model-based boosting (mboost, Hofner et al. (2014)) with a logistic classification model for the prediction of missing ratings. This method was preferred over other classification models for four main reasons: the embedding of variable selection, good handling of multicollinearity, the possibility to include weights, and interpretability. During estimation, the algorithm carries out variable and model selection, meaning that only relevant therapies affect the prediction. Moreover, mboost is known to perform well in cases of multicollinearity, which is likely to be present in health data, where specific values depend on one another. Furthermore, observation weights can be incorporated into the component-wise gradient boosting process for weighted regression. This is particularly useful in the case of imbalanced classes, since assigning weights to all observations gives appropriate importance to observations from the majority and minority class. Finally, the results, including effect estimates, can be interpreted in the same way as in common statistical model estimates (Hofner et al., 2014). This is a particularly important in the considered context because of the need to understand how the decision-making process is influenced.

##### Approach IIb: Hybrid model-based HRS integrating patient covariates

As an extension of Approach IIa, we additionally incorporate patient core data as input values into the classification models of the HRS. Consequently, patient data is embedded not only indirectly through the initial values derived from memory-based techniques, but also directly affecting the prediction of therapy ratings. In this approach, the variable selection process, which is implemented by the mboost algorithm, is of particular relevance, given the increasing number of covariates that arise when additional patient data is incorporated.

#### 2.4.3 Evaluation of RS

RS generally aim at several desirable characteristics, such as coverage, novelty, serendipity, utility, adaptivity, and scalability. Prediction accuracy is one of the most important objectives (Gunawardana et al., 2022). In line with this, the overarching objective of our HRS is to predict therapy response. In doing so, we focus on patients who have newly been diagnosed with sepsis. Naturally, at the time of diagnosis, there is no response data (for no therapy) available for the patient of interest.

In Section 2.4.4, we will implement the four proposed HRS using real data. Our interest lies in determining which HRS best meets the formulated prediction objective. For this reason, we address the question of evaluation on the matrix completion level (rather than the recommendation level). To avoid over- or underfitting, we distinguish between training and evaluation data when evaluating a system’s performance. The patient-therapy matrix is completed based on the training data, that is the patient-therapy matrix after removal of observed values from the evaluation set. This results in a complete patient-therapy matrix with predicted values for unobserved or removed ratings. To measure performance, the predictions are compared to the entries of the evaluation set where applicable.

The partitioning into training and evaluation set is done in two different ways (both applied in our study), illustrated in Figure 4:

- **Row-wise splitting:** As in the classical prediction context, we randomly select a previously specified proportion of patients (that is, whole rows) to belong to the evaluation set. The resulting training data does not contain any therapy rating for these patients at all.
- **Cell-wise splitting:** We randomly select single (observed) entries of the patient-therapy matrix (instead of whole rows) to belong to the evaluation set such that a previously specified proportion of observed values is used for evaluation (Aggarwal, 2016).

**Figure 4:**
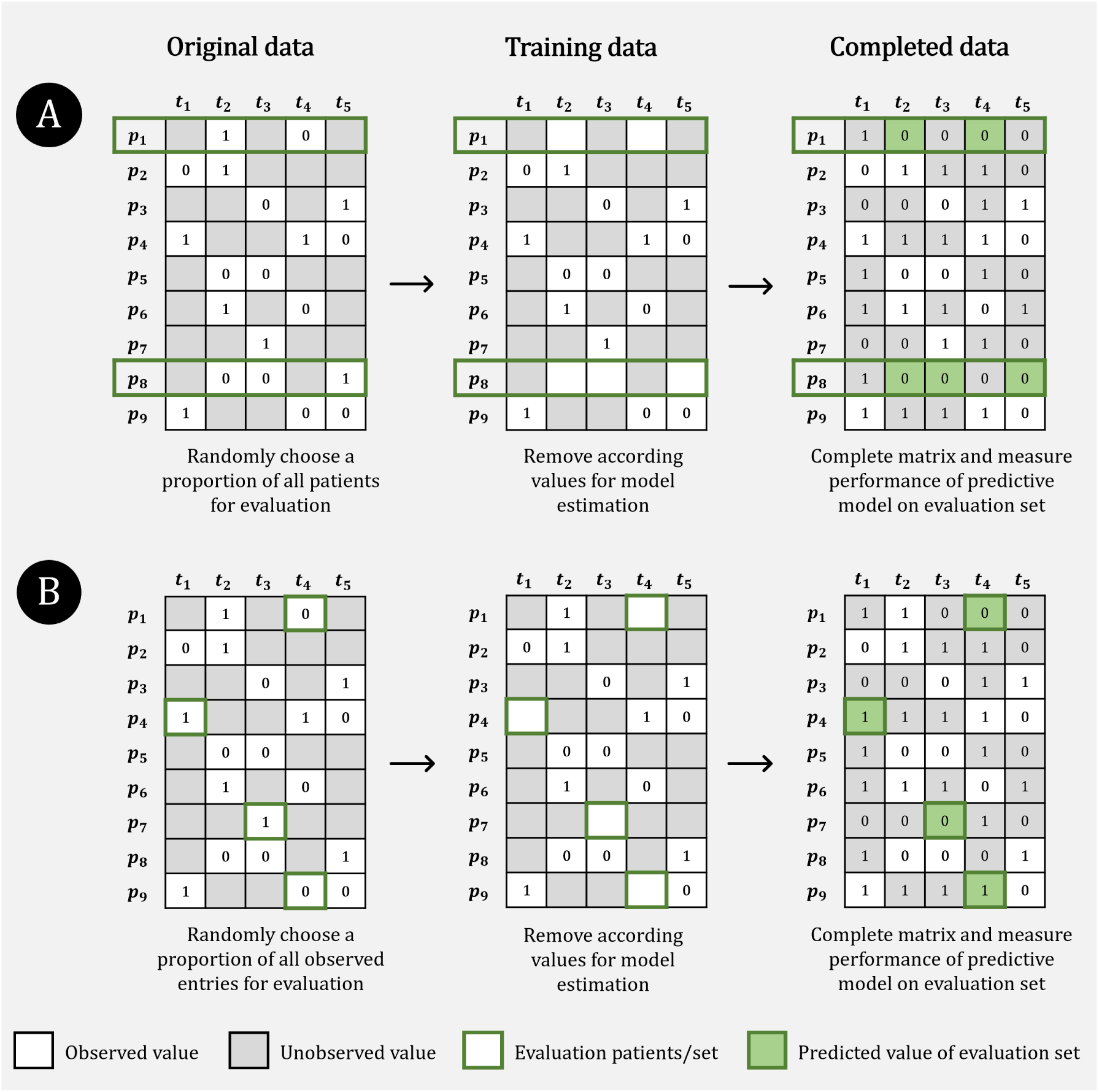
Splitting strategies for evaluation of matrix completion in HRS. **A. Row-wise splitting:** Complete observations (patients) are used as evaluation data and observed data is removed for model estimation; after completion of the whole patient-therapy matrix, evaluation is done on originally observed values from patients in evaluation set. **B. Cell-wise splitting:** Single entries of the whole matrix are removed for model estimation, and originally observed values are used as evaluation set after prediction.

Each splitting strategy has its merits and is suited to distinct scenarios. The cell-wise splitting strategy corresponds to the use case where some therapies are already rated for the patients of interest, while no information about therapies is available in row-wise splitting (cold start problem). While we expect comparable performance results for both splitting strategies when applied in the context of memory-based HRS, we anticipate poorer results with row-wise splitting when evaluating model-based HRS. This expectation results from the different ways the data is taken into account: in the memory-based HRS, only data from similar patients is included for prediction, whereas the model-based HRS include all observations.

#### 2.4.4 Application on sepsis data

All four presented HRS approaches are applied to the data introduced in Section 2.1 with the aim of completing the patient-therapy matrix ***R***, including predictions for new patients.

The memory-based HRS Ia and Ib require calculations of similarities for patients and therapies. We first identify sets of similar patients, once by setting various numbers of similar patients *ν* ∈ {10, 50, 100, 250, 500, 1000}, once by setting various patient similarity thresholds, *ρ* ∈ {0.95, 0.90, *…,* 0.60} (in steps of 0.05). Based on these, we determine sets of similar therapies for similar patients for various therapy similarity thresholds, *τ* ∈ {1.00, 0.95, *…,* 0.70} (in steps of 0.05).

For the two model-based HRS IIa and IIb, we apply both Approach Ia and Approach Ib for initializing the originally incomplete matrix ***R***. For the subsequent iterative procedure, we employ a logistic model-based boosting algorithm (mboost, see Section 2.4). All variables (therapies and, in IIb, patient covariates) are added as linear base-learners to the model. The tuning parameters include the initial number of boosting iterations mstop = 500 and a step length parameter of 0.05. In order to choose an appropriate number of boosting iterations, the empirical risk for different stopping parameters is estimated and optimized by 10-fold cross-validation (CV) as suggested by Hofner et al. (2014). By optimizing the stopping criterion, we select the most influential predictors. For matrix completion, we apply both presented ways of updating the entries of the rating matrix, collective and immediate, and stop after 50 iterations. For Approach IIb, variables from patient core data as well as blood values and vital signs (listed in Table 1) are incorporated as additional covariates for the classification model.

For model evaluation and comparison, we apply both presented splitting strategies (row-wise and cell-wise) to each HRS approach. For each HRS, we perform ten-fold CV (for model-based HRS nested CV) (Krstajic et al., 2014). To compare the approaches, we use the performance measures AUC, sensitivity, specificity, precision and F1 score, using a classification threshold of 0.5. The final measures represent the mean performance calculated across all ten CV folds. Given the HRS’s objective of identifying suitable therapies for sepsis patients, it is crucial to ensure that the recommended therapies are truly effective. If therapies predicted to yield a high response are, in fact, effective, the evaluation of alternative therapies becomes less important. Therefore, we mainly focus on precision and F1 score. Precision represents the ratio between the number of correctly positive classified cases with respect to all cases classified as positive. The F1 score is the harmonic mean of sensitivity and precision, and is particularly useful if class imbalance is present in the target variable (Saito and Rehmsmeier, 2015).

The results obtained from this application are presented in Section 3, where the various HRS approaches are compared among each other with regard to their performance and relevance for clinical use.

## 3 Results

The performance of an HRS inevitably depends on the underlying data and is, whether explicitly or implicitly, influenced by the similarity among patients as well as among therapies. The pairwise calculated patient similarities according to Equation (1) range from 0.12 to 0.996 (mean 0.76, median 0.76), excluding a patient’s similarity with themselves. A heatmap of all patient similarities is shown in Figure A4. The calculation of pairwise therapy similarities as defined in Equation (5) requires information about pathogen coverage and pathogen-specific therapy similarity, described by Equation (4). Figure 5 displays the pathogen-therapy matrix that is used for our analysis. Some therapies cover only a small, others a larger number of pathogens; some therapies cover identical set of pathogens. The resulting therapy similarities (excluding self-similarities) range from 0.13 to 1.00 (mean 0.57, median 0.56). Figure A5 shows a heatmap of the pairwise similarities for all therapies that are included in our study.

**Figure 5:**
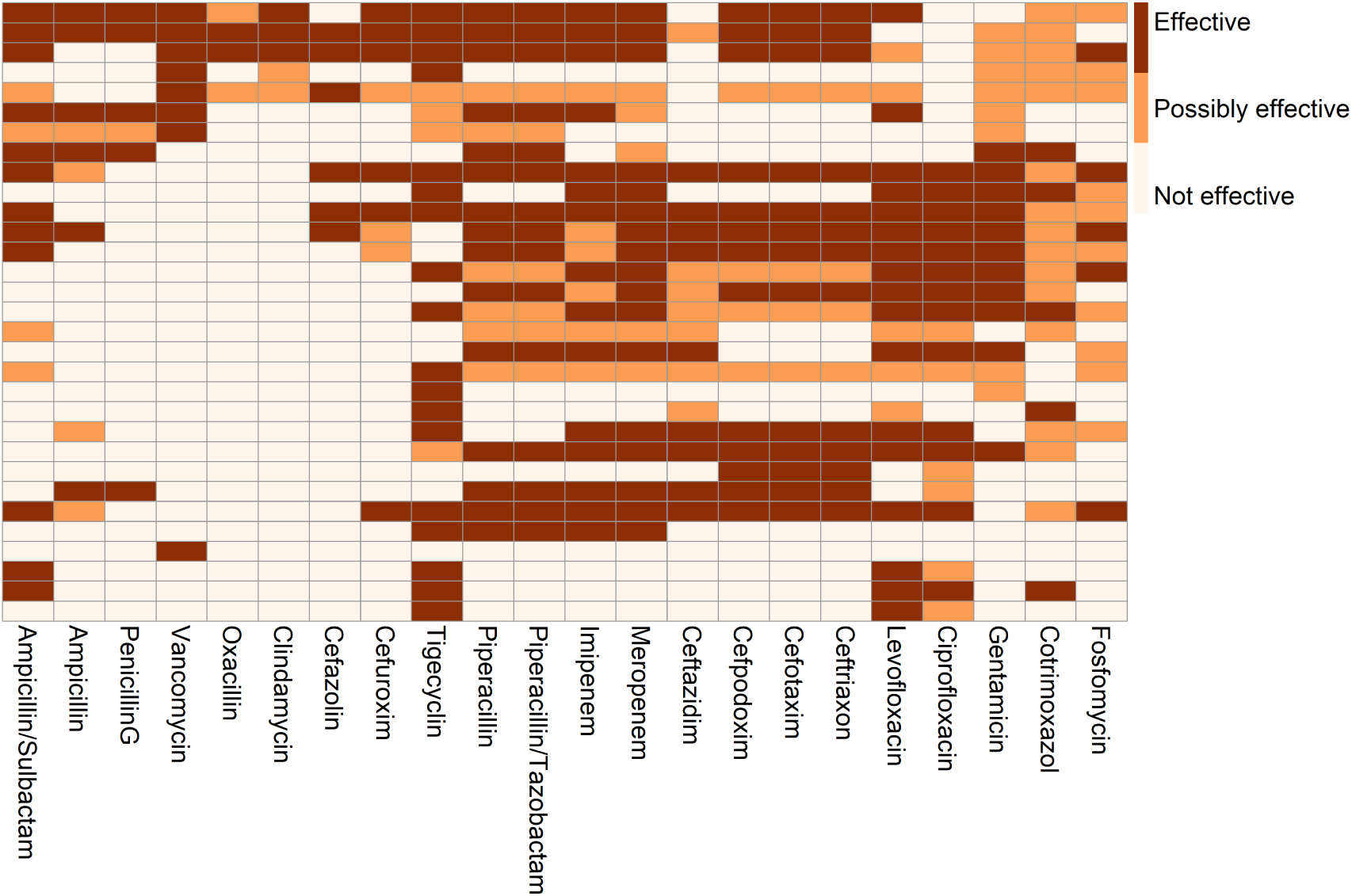
Pathogen-therapy matrix for the 31 pathogens and 22 therapies used for building the HRS. Pathogens are shown in rows, therapies in columns. The color of each cell depicts the general coverage of a therapy against the respective pathogen as explained in the context of Equation (3).

Section 2.4 introduced four fundamental approaches to HRS: two memory-based methods (Ia and Ib) and two model-based methods (IIa and IIb). Within each approach, several design choices arose — for instance, in memory-based methods, the selection of similarity thresholds, and in model-based methods, the choice of initialization and update schemes. For the evaluation, both the splitting scheme and the evaluation measure are of importance. In the following, we compare not only the four HRS approaches with each other but also examine, within each approach, the impact of choosing appropriate parameter settings. In addition to selected individual results, we present general trends observed.

### Approach Ia

Performance results for Approach Ia are shown in Table A2, considering various choices of similarity thresholds *ν* and *ρ*, and both splitting strategies. The best outcomes in terms of F1 score are achieved with a patient similarity threshold of *ν* = 100 and *ρ* = 0.8, respectively (the threshold of *ρ* = 0.8 results in an average of 213 patients being recognized as similar per patient). These values appear to mark an optimal point for our data in the sense that the F1 score increases (almost) consistently with growing *ν* until it reaches its maximum, and then decreases (almost) consistently as *ν* continues to increase. The same pattern can be observed for the influence of *ρ*, where a larger *ρ* corresponds to a smaller *ν*. The choice of splitting scheme hardly affects the performance in Approach Ia. Overall, the performance is satisfyingly high for AUC, sensitivity, precision and F1 score (AUC and precision around 0.75, sensitivity around 0.89, and F1 around 0.82), while the specificity is comparably low (around 0.42). This indicates that susceptible therapies are likely to be correctly identified, while truly resistant therapies are detected with only low probability.

### Approach Ib

Performance results from Approach Ib are shown in Table A3. Based on the analysis for Approach Ia, patient similarity thresholds were set to *ν* = 100 and *ρ* = 0.8, respectively, and we explore the outcomes for different values of the therapy similarity threshold *τ* and the application of each splitting strategy. With respect to F1 score, best results are achieved for therapy similarity thresholds *τ* ≥ 0.9. Notably, they differ within *τ* ∈ {0.9, 0.95, 1.00} only regarding AUC. Setting the threshold to *τ* = 1.00 means that only therapies with identical sets of covered pathogens would be used for estimation of ratings, which is close (but not identical to) Approach Ia. To better explore the additional potential of Approach Ib, we use the threshold *τ* = 0.90 to also include therapies covering a similar but not identical set of pathogens. As for Approach Ia, the choice of splitting scheme has negligible impact. Furthermore, AUC, sensitivity, precision and F1 score are high, with sensitivity now even ranging from 0.904 to 0.975. As in Approach Ia, however, specificity is again low, now around 0.3 to 0.34. The already observed discrepancy between sensitivity and specificity is therefore even more pronounced here.

### Approaches IIa and IIb

Table A4 shows the performance results for Approaches IIa and IIb for various combinations of splitting scheme, initialization scheme, and matrix updates. In particular, both Approach Ia and Ib are used for initialization of the patient-therapy matrix. In doing so, based on the results described above, the patient similarity threshold is always set to *ρ* = 0.8, the therapy similarity threshold to *τ* = 0.90. Other than for Approaches Ia and Ib, the performance changes notably between the two splitting strategies, with considerably better values for cell-wise splitting. In contrast, the performance regarding collective and immediate matrix updates do not differ substantially. Also, using either Approach Ia or Ib for initialization does not greatly affect the results.

While the results presented so far focused on performance measures, the model-based Approaches IIa and IIb offer an additional advantage: they allow an examination of which variables ultimately contributed to the estimation of ratings and, consequently, to recommendations of therapies. In our implementation, namely, the application of mboost, it is possible (as for several other classifiers) to identify which variables were selected by the respective model as covariates for predicting the therapy response of specific antibiotic therapies. The strength of the effect on the target variable can be determined from the respective model coefficients. This provides further insight into the relationship between therapies and other influencing factors. Figures A6 and A7 graphically present the chosen covariates for an exemplary set of model specifications.

To summarize the identified choices leading to optimal performance per approach and splitting strategy, Table 3 displays both these settings and the resulting scores. A comparison across the approaches yields the following: Overall, regarding F1 score, we achieve the best result of a memory-based HRS with Approach Ia for row-wise splitting, and for a model-based HRS with Approach IIb for cell-wise splitting. Differences in F1 score between Approach Ia and Ib and between Approach IIa and IIb are negligible, often only visible to the second decimal place. With row-wise splitting, memory-based methods produce notably better AUC, sensitivity, precision and F1 score than model-based methods, but worse specificity. With cell-wise splitting, in contrast, the model-based approaches perform better regarding AUC, specificity, precision and F1 score than the memory-based approaches. Thus, regarding specificity, the model-based approaches outperform the memory-based ones independently of the splitting scheme. The overall best F1 score is reached by Approach IIb.

**Table 3:**
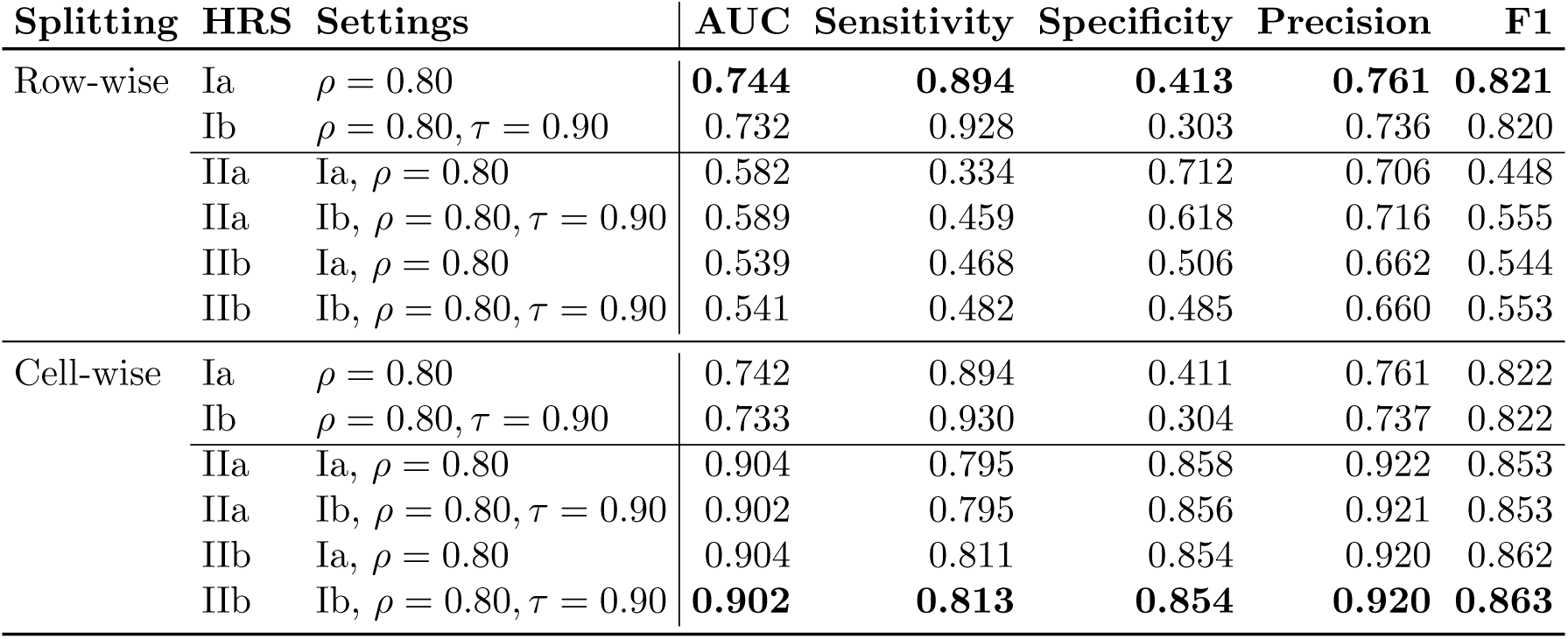
Results of matrix completion in HRS approaches: Performance measures AUC, sensitivity, specificity, precision and F1 score are shown for various combinations of splitting strategy, HRS approach, similarity thresholds and initialization scheme in model-based matrix completion with immediate matrix update. Best results regarding F1 score for each splitting strategy are highlighted in bold.

While the comparison so far considers the performance of the approaches across all therapies, an analysis at the level of individual therapies reveals a different pattern: Table 4 displays the sensitivity, specificity and F1 score for each therapy separately (mean over ten CV folds). Notably, for Approaches Ia and Ib, many therapies show either a sensitivity of one and specificity of zero, or vice versa. In contrast, the model-based Approaches IIa and IIb demonstrate a more balanced trade-off between sensitivity and specificity across therapies. This difference between the approaches is even more evident for cell-wise splitting (see Table A5). Overall, the optimized model-based approaches show a clearly superior performance over the best memory-based ones, both with respect to single therapies as well as regarding the overall picture.

**Table 4:**
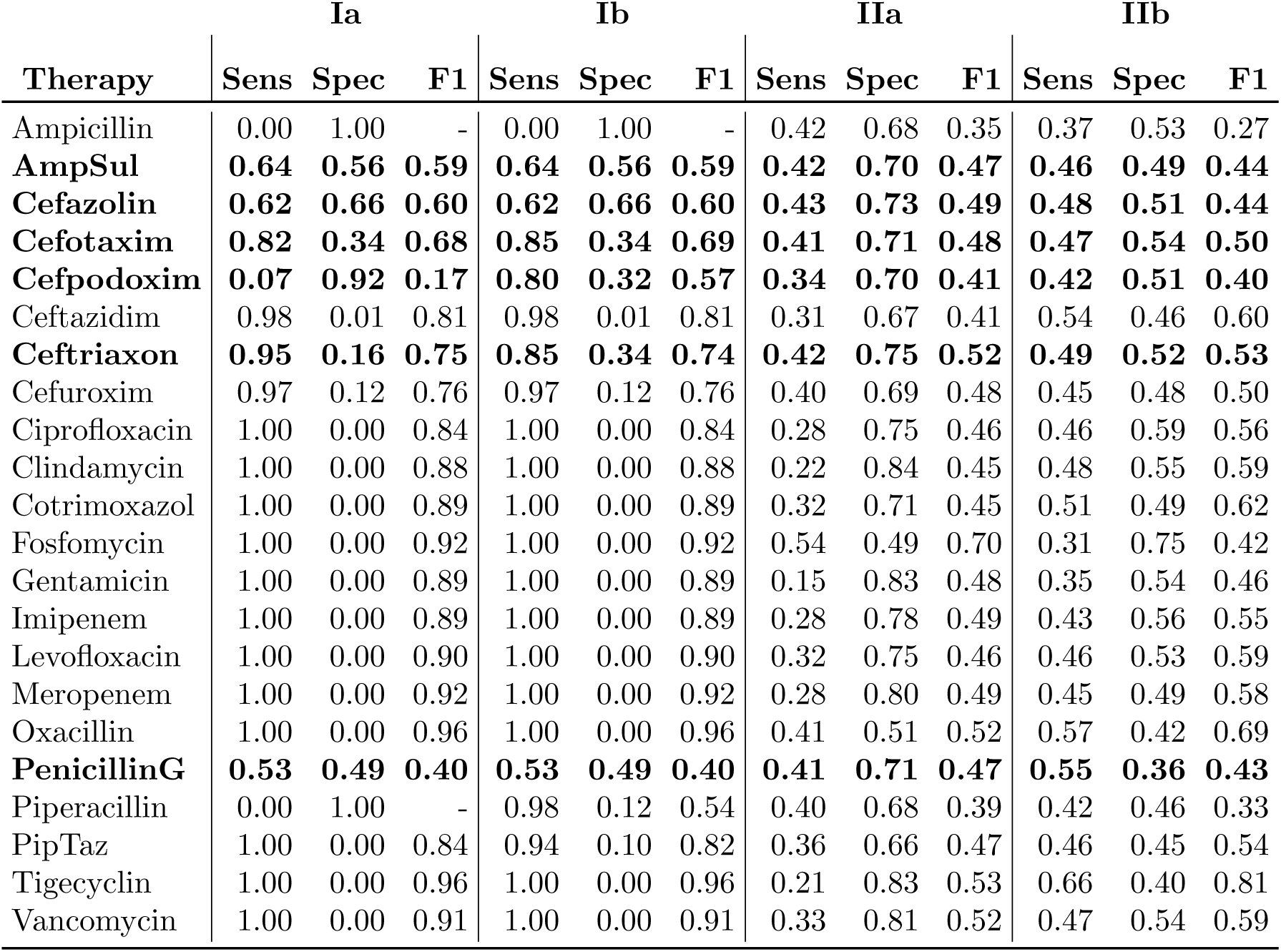
Therapy-specific performance of matrix completion in the four HRS approaches: Mean sensitivity, specificity and F1 score across all CV folds. Since the F1 score is particularly useful in case of class imbalance (with respect to observed resistance statuses ‘S’ and ‘R’ per therapy), therapies with a rather balanced resistance class ratio between 0.66 and 1.5 are printed in bold. Where no F1 value is shown, calculation was impossible because of a denominator equal to zero. Results displayed here are based on row-wise splitting; respective results for cell-wise splitting can be found in Table A5.

|  | Ia |  |  | Ib |  |  | IIa |  |  | IIb |  |  |
| --- | --- | --- | --- | --- | --- | --- | --- | --- | --- | --- | --- | --- |
| Therapy | Sens | Spec | F1 | Sens | Spec | F1 | Sens | Spec | F1 | Sens | Spec | F1 |
| Ampicillin | 0.00 | 1.00 | - | 0.00 | 1.00 | - | 0.42 | 0.68 | 0.35 | 0.37 | 0.53 | 0.27 |
| **AmpSul** | **0.64** | **0.56** | **0.59** | **0.64** | **0.56** | **0.59** | **0.42** | **0.70** | **0.47** | **0.46** | **0.49** | **0.44** |
| **Cefazolin** | **0.62** | **0.66** | **0.60** | **0.62** | **0.66** | **0.60** | **0.43** | **0.73** | **0.49** | **0.48** | **0.51** | **0.44** |
| **Cefotaxim** | **0.82** | **0.34** | **0.68** | **0.85** | **0.34** | **0.69** | **0.41** | **0.71** | **0.48** | **0.47** | **0.54** | **0.50** |
| **Cefpodoxim** | **0.07** | **0.92** | **0.17** | **0.80** | **0.32** | **0.57** | **0.34** | **0.70** | **0.41** | **0.42** | **0.51** | **0.40** |
| Ceftazidim | 0.98 | 0.01 | 0.81 | 0.98 | 0.01 | 0.81 | 0.31 | 0.67 | 0.41 | 0.54 | 0.46 | 0.60 |
| **Ceftriaxon** | **0.95** | **0.16** | **0.75** | **0.85** | **0.34** | **0.74** | **0.42** | **0.75** | **0.52** | **0.49** | **0.52** | **0.53** |
| Cefuroxim | 0.97 | 0.12 | 0.76 | 0.97 | 0.12 | 0.76 | 0.40 | 0.69 | 0.48 | 0.45 | 0.48 | 0.50 |
| Ciprofloxacin | 1.00 | 0.00 | 0.84 | 1.00 | 0.00 | 0.84 | 0.28 | 0.75 | 0.46 | 0.46 | 0.59 | 0.56 |
| Clindamycin | 1.00 | 0.00 | 0.88 | 1.00 | 0.00 | 0.88 | 0.22 | 0.84 | 0.45 | 0.48 | 0.55 | 0.59 |
| Cotrimoxazol | 1.00 | 0.00 | 0.89 | 1.00 | 0.00 | 0.89 | 0.32 | 0.71 | 0.45 | 0.51 | 0.49 | 0.62 |
| Fosfomycin | 1.00 | 0.00 | 0.92 | 1.00 | 0.00 | 0.92 | 0.54 | 0.49 | 0.70 | 0.31 | 0.75 | 0.42 |
| Gentamicin | 1.00 | 0.00 | 0.89 | 1.00 | 0.00 | 0.89 | 0.15 | 0.83 | 0.48 | 0.35 | 0.54 | 0.46 |
| Imipenem | 1.00 | 0.00 | 0.89 | 1.00 | 0.00 | 0.89 | 0.28 | 0.78 | 0.49 | 0.43 | 0.56 | 0.55 |
| Levofloxacin | 1.00 | 0.00 | 0.90 | 1.00 | 0.00 | 0.90 | 0.32 | 0.75 | 0.46 | 0.46 | 0.53 | 0.59 |
| Meropenem | 1.00 | 0.00 | 0.92 | 1.00 | 0.00 | 0.92 | 0.28 | 0.80 | 0.49 | 0.45 | 0.49 | 0.58 |
| Oxacillin | 1.00 | 0.00 | 0.96 | 1.00 | 0.00 | 0.96 | 0.41 | 0.51 | 0.52 | 0.57 | 0.42 | 0.69 |
| **PenicillinG** | **0.53** | **0.49** | **0.40** | **0.53** | **0.49** | **0.40** | **0.41** | **0.71** | **0.47** | **0.55** | **0.36** | **0.43** |
| Piperacillin | 0.00 | 1.00 | - | 0.98 | 0.12 | 0.54 | 0.40 | 0.68 | 0.39 | 0.42 | 0.46 | 0.33 |
| PipTaz | 1.00 | 0.00 | 0.84 | 0.94 | 0.10 | 0.82 | 0.36 | 0.66 | 0.47 | 0.46 | 0.45 | 0.54 |
| Tigecyclin | 1.00 | 0.00 | 0.96 | 1.00 | 0.00 | 0.96 | 0.21 | 0.83 | 0.53 | 0.66 | 0.40 | 0.81 |
| Vancomycin | 1.00 | 0.00 | 0.91 | 1.00 | 0.00 | 0.91 | 0.33 | 0.81 | 0.52 | 0.47 | 0.54 | 0.59 |

## 4 Discussion

The potential of HRS is shaped, on the one hand, by the wide range of available methods originating from diverse contexts far beyond the healthcare domain. At the same time, they must pay close attention to context, data, guidelines, and other domain-specific factors. Using the example of initial antibiotic administration for newly diagnosed sepsis patients, we have explored — based on a concrete dataset — how different methodological approaches can be applied, what levels of performance they achieve, and which additional strengths and limitations they exhibit.

### Strengths and weaknesses of proposed HRS

The superior performance of the optimized model-based approaches with cell-wise splitting, observed in Section 3, suggests that predictions for unrated therapies generally benefit from ratings that were available for the same patient but different therapies. One may thus hypothesize that prediction could be substantially improved if ratings were available for even more therapies (e. g. information about antibiotics that patients received before being admitted to the ICU).

Restricting the number of patients regarded as sufficiently similar in the two memory-based approaches enhances the personalization of recommendations. However, this restriction simultaneously reduces the amount of information considered for individual therapies and limits variation. For our sepsis patient dataset, a patient similarity threshold of *ρ* = 0.8 appears to provide the right balance between patient similarity and an adequate sample size for reliable predictions. However, the optimal threshold will vary depending on the size, quality and comprehensiveness of the dataset and should therefore be determined for each new analysis.

The performance results for Approach Ib suggest that incorporating information from similar therapies does not generally improve the predictive performance compared to Approach Ia. Although including similar therapies increases the number of observed ratings available for prediction, it may also introduce uncertainty by weighing ratings with both patient and therapy similarity. Despite this, Approach Ib may still be advantageous in scenarios where the similarity structure among therapies is different.

The model-based Approach IIb integrates additional patient covariates to the otherwise identical Approach IIa. It outperforms Approach IIa, but not notably. This indicates that clinical variables such as core data, blood values and vital signs do not influence therapy resistance to a large extent, and that resistance mainly depends on the present pathogens. Another explanation is that our data misses relevant covariates. As discussed in Schmiegel et al. (2025), additional variables such as the patient’s origin, travel history, profession, previous administration of antibiotics and local resistances are likely to influence resistance to specific therapies.

We observed considerable variation in the performance regarding different individual therapies (see Table 4 and Table A5), particularly in Approaches Ia and Ib. There are several cases with a sensitivity of one and specificity of zero or vice versa; in these cases, the model is exclusively predicting one same class (i. e. always ‘susceptible’ or always ‘resistant’) for the corresponding therapy. This is a frequently observed phenomenon in the context of highly imbalanced datasets, where the model tends to favor the majority class. Thus, in our case, the variation is likely attributable to the substantial imbalance in resistance classes among certain therapies. Furthermore, these extreme values can result from the absence of one class in the evaluation set, in which case the corresponding metric (sensitivity or specificity) is not meaningful. Approaches Ia and Ib do not allow for class rebalancing or the inclusion of class weights. Consequently, therapies with a high class imbalance (i. e., a class ratio less than or equal to 0.66) and ‘susceptible’ as majority class show high sensitivity but low specificity, and vice versa with ‘resistant’ as majority class. In contrast, therapies with more balanced class distributions, such as Ampicillin/Sulbactam or Cefazolin, yield similar results for sensitivity and specificity. The amount of unbalanced therapies contributes to the elevated overall F1 scores in Approaches Ia and Ib (see Table 3). Approaches IIa and IIb, incorporating weighted regression, show less variation across therapies and a more balanced relationship between sensitivity and specificity compared to memory-based approaches. While they yield lower F1 scores, they address class imbalance more effectively and thus provide more robust and reliable results.

A further advantage of Approaches IIa and IIb is the enhanced interpretability and explainability due to the use of a classification model such as mboost. Through variable selection, the relevant influencing factors for the corresponding therapy can be identified, and the strength of their effect can be interpreted through the model coefficients.

Compared to memory-based methods, model-based approaches are more time-consuming and computationally expensive, especially if one chooses a strict interpretation of when convergence is reached. Furthermore, the model-based approaches presented here use one of the memory-based approaches as input values, thus summing up the effort of two approaches. A straightforward alternative initialization such as using plain mean values would not be appropriate for the considered application, as every newly added patient would receive the same initial values and therefore receive the same prediction.

Approaches Ia, Ib and IIa have the advantage that they can handle missing values in patient data whereas Approach IIb requires complete data (used as model covariates), which may demand imputation of missing data. Since Approach Ib incorporates information about similar therapies, this is the only approach that can make predictions if there are no similar patients available with ratings of the therapy of interest, or when new therapies are introduced for which there are no patient ratings yet.

### Data quality, bias and uncertainty

As in any data analysis, certain characteristics of the data can substantially influence the results. The already described substantial class imbalance for some of the therapies can lead to unsatisfactory performance if not addressed. A countermeasure lies e. g. in applying weights. However, including the imbalanced therapies as covariates in Approaches IIa and IIb may negatively impact the predictions and lead to biased estimates.

The number of patients in our application is tiny compared to several everyday RS use cases (e. g., compared to users of shopping platforms). A larger sample size would most likely result in more distinct patient groups in terms of similarity, provide more detailed information about each therapy and lead to more precise predictions. Further, a larger sample size would offer more possibilities for data rebalancing. However, since small sample sizes are common in the medical context, we have chosen HRS methods that work well with a relatively small number of patients. The problem of small datasets is a common issue in the health context. It can be addressed by combining multiple datasets; however, this in turn introduces other challenges. Düsing et al. (2024), for example, describe the complexity of federated learning as a possible approach to overcoming the issue of small sample sizes in case of health record data.

Our data comes with uncertainty for several reasons: AST results may change over time for specific pathogens; we consider only one time point. Furthermore, in order to obtain one resistance result per patient and therapy, we need to summarize the results across all present pathogens; it is not possible to determine which pathogen is causing the sepsis. Additionally, the patient-therapy matrix contains a systemic bias, as the therapies to be tested with AST are not randomly selected for each patient, but are instead chosen in advance by the treating physicians or laboratory staff. An alternative to using AST data would be to use true information about the effectiveness of therapies that have actually been administered, ideally in a numeric rather than categorical format. However, our data does not contain any information about therapy effectiveness. From a quantitative (i. e. data-based) perspective, it is challenging to determine the effectiveness of a specific therapy that was given to a patient. Furthermore, each patient is administered only a small number of therapies, resulting in a patient-therapy matrix that is even sparser than with AST data.

As outlined in Schmiegel et al. (2025), using alternative data sources such as the MIMIC IV database (Medical Information Mart for Intensive Care, Johnson et al. (2023)) for application at a specific site or region is problematic, as important information is often missing and guidelines are not necessarily comparable across different countries.

## Methods

The use of a hybrid approach for our application enhances the ability to predict therapy responses for newly diagnosed patients. The system’s capacity to identify previously unknown patterns or influencing factors is noteworthy, as it depends not solely on predefined guidelines and existing information on therapy responses. The system is characterized by a high level of flexibility, achieved through the integration of multiple algorithms in a hybrid manner, and is capable of scaling to accommodate even large sample sizes. Furthermore, HRS can continuously adapt, incorporating information from rarely administered or newly introduced therapies.

Overall, the memory-based Approaches Ia and Ib depend strongly on patient and therapy similarity. Both approaches could be improved by using different variable weights to calculate Gower’s similarity (or e. g. using feature weighting algorithms as in Gräßer et al. (2017) and Gräßer et al. (2019)), or even by using different definitions of similarity. An alternative method of identifying similar patients is cluster analysis. This technique involves assigning a new patient to one of several predefined groups. Seymour et al. (2019) identified four clinical phenotypes that could be used as predefined patient subgroups. However, relying solely on data from patient health records available at the time of diagnosis, as in our case, may render these four subgroups insufficient for accounting for individual patient variations. By employing the proposed similarity coefficient, we aim to maintain a more patient-specific analysis that is as flexible and tailored as possible.

Regarding Approaches IIa and IIb, it would be possible to apply alternative classifiers such as random forest, support vector machines or extreme gradient boosting. However, we do not expect that these will demonstrate a substantial improvement in performance. Moreover, the interpretability of these would be inferior to that of mboost, in which the coefficients of the model can be interpreted directly.

### Integration in CDSS

When incorporating an HRS into a CDSS for sepsis management, the process could be as follows: The HRS predicts the expected therapy response for each patient-therapy combination. The therapies can then be ranked for each patient according to their predicted response. Within the CDSS, the highest-ranked therapies would be recommended. The physician is then responsible for selecting the most appropriate therapy, taking into account risk factors such as comorbidities, allergies, and other limiting medical conditions. Tokgöz et al. (2024) examine relevant factors for the implementation of AI-based CDSS for antibiotic prescription in hospitals from the perspective of hospital managers.

The most important limitation of HRS with regard to generalizability is that, in practice, a separate system needs to be built for each hospital. This is because the underlying data is not comparable due to diverse processes for data documentation, different guidelines and varying administration for certain therapies.

## 5 Conclusion

Sepsis is a life-threatening condition that requires rapid treatment to increase the chance of survival. To address the research question of finding the most suitable therapy for sepsis patients at the time of diagnosis, we developed various hybrid HRS approaches. These approaches are designed to be integrated into a comprehensive CDSS for sepsis management, with the ultimate goal of improving treatment decisions.

The HRS presented are suitable for different scenarios: Memory-based methods seem to be more suitable for new patients without any therapy information if therapy data is balanced, while model-based approaches obtain better results in case that some values for therapy response are already present for new patients. Overall, however, the results indicate that, based on the current data, the HRS approaches are still in an early stage of development and offer potential for further improvement before clinical application. We assume that the primary factors contributing to this are the dataset used for model training and the considerable individuality among patients. This is in line with Vincent (2025) who states that ‘patients with sepsis are so different in terms of demographics, co-morbidities, genetics, causative microorganisms, stage of disease at presentation, prior treatments, and degree of host response, among other factors, that identifying a single agent that would work for all was never going to be realistic.’ Consequently, we emphasize the importance of high-quality data that meets specific criteria.

Despite these limitations, we have outlined a methodological framework for HRS applicable to various scenarios, which can serve as a foundation for future refinement and extension. Sepsis therapy remains a highly relevant field that requires continued research to achieve tangible benefits for future patients.

## Data Availability

Due to data privacy protection, we are not allowed to share the data of the German hospital used in this work.

https://github.com/fuchslab/Hybrid_HRS_Sepsis

## List of abbreviations

AI: Artificial intelligence
AMR: Antimicrobial resistance
AST: Antimicrobial susceptibility testing
CBF: Content-based filtering
CDSS: Clinical decision support system
CF: Collaborative filtering
CV: Cross-validation
DSS: Decision support system
HRS: Health recommender system
ICU: Intensive care unit
KRP: Top-k recommendation problem
MCP: Matrix completion problem
mboost: Model-based boosting
ML: Machine learning
RS: Recommender system

## Computational details

The data preprocessing and analysis was done using the statistical software R version 4.4.3 (R Core Team, 2025). The code used for analysis is available at https://github.com/fuchslab/Hybrid_HRS_Sepsis.

## Declarations

### Ethics approval and consent to participate

Ethical approval was given by the Ethics Committee of the Westphalia-Lippe Medical Association and the University of Münster (Ethik-Kommission der Ärztekammer Westfalen-Lippe und der Westfälischen Wilhelms-Universität Münster); case number: 2021-699-f-S.

### Consent for publication

Not applicable.

### Authors’ contributions

H.M., C.F. and S.S. initiated the project and contributed to the conceptualization of the research goals. R.B. and S.R. provided the data of sepsis patients, gave input on clinical aspects and assured realistic assumptions. The data preprocessing was carried out jointly by H.M. and S.S. H.M. conducted literature research, method development and implementation, data analysis and visualization, and drafted the manuscript. H.M., C.F. and S.S. edited and critically revised the manuscript. All authors commented on the manuscript and approved the final version.

## Acknowledgements

We would like to thank Tamara Schamberger and Gregor Miller for helpful feedback and discussions. Further, we thank all members of the ‘KINBIOTICS’ project.

## A Supplementary Material

### A.1 Data Preprocessing

The study involves extensive data preprocessing, which could only be outlined briefly in the main manuscript for the sake of conciseness. In the interest of transparency and reproducibility, more detailed descriptions are provided here.

#### A.1.1 Main dataset

##### Categorization and missing values

The suspected infection focus is included as categorical variable. We summarize those suspected infection focuses occurring in fewer than 50 cases as ‘other’. Besides, we remove variables from patients’ core data and laboratory measurements which contain more than 35 % missing values. Furthermore, we remove the variable for SOFA score since it is composed by the following variables which are already contained in the data: invasive mean blood pressure, fraction of inspired oxygen, pressure of oxygen, glomerular filtration rate, bilirubin, thrombocytes, creatinine. Figure A1 shows the missingness pattern after this preprocessing.

**Figure A1:**
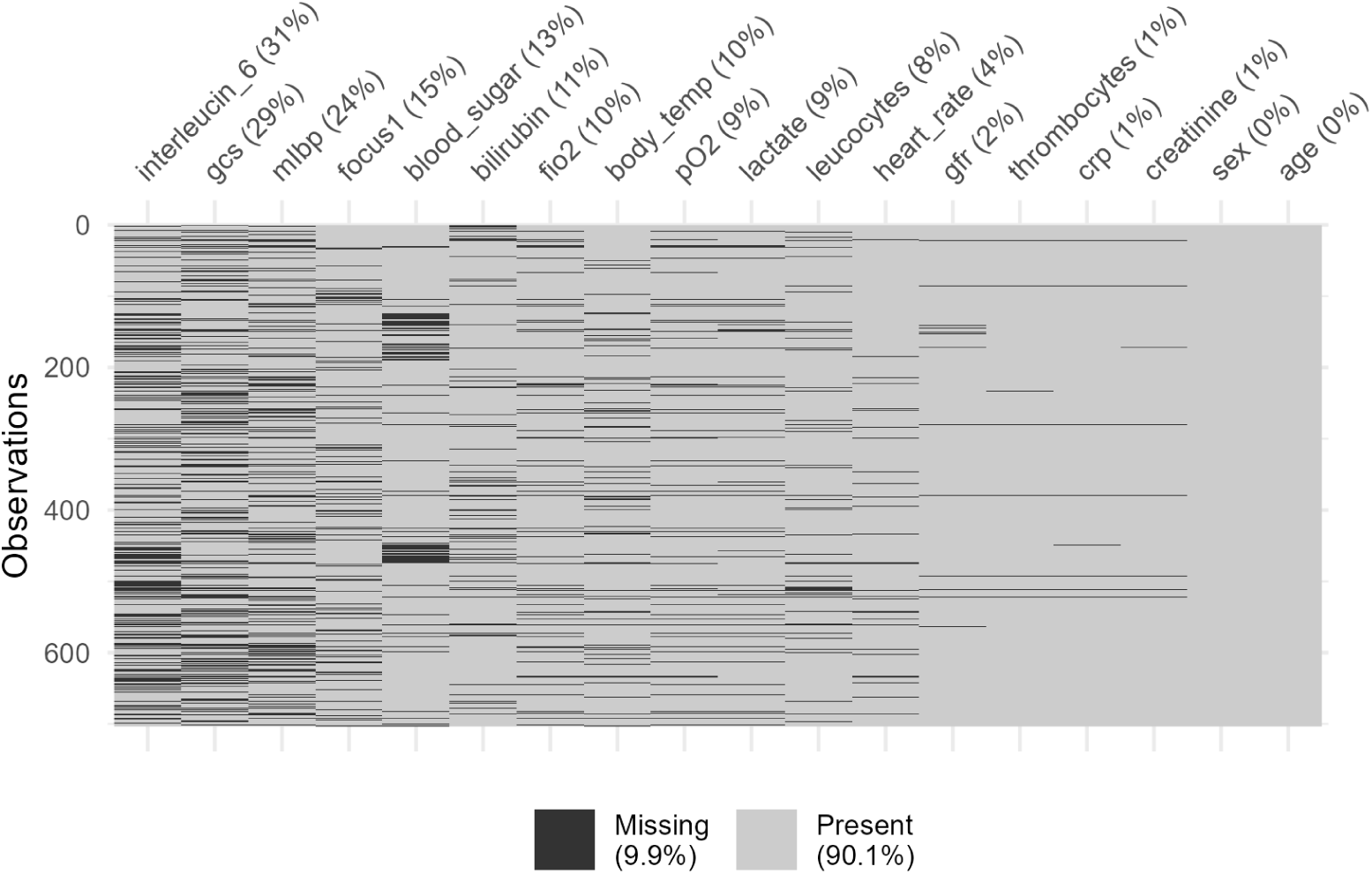
Percentages and pattern of missing values in variables of the final dataset. Observations (infection IDs) are shown in rows, variables in columns. Black color marks missing values while gray color reflects observed entries.

##### Antimicrobial susceptibility testing

For data from AST, the following rule is applied: only results of testings are considered which were ordered either on the same day as the first antibiotic treatment or at maximum 24 hours previous to it. Further, we keep only results where pathogens were tested positively with high load (apart from the pathogens ‘Candida’ in blood samples and ‘Aspergillus’ for which we accepted also sparse occurrence). Following the new definitions of susceptibility categories EUCAST 2019 (Nabal Díaz et al., 2022), we set all AST results with outcome ‘I (intermediate)’ to ‘S (susceptible)’ and consequently keep only two categories (susceptible and resistant).

Since it is also possible to administer combined antibiotic therapies, we additionally construct AST variables for the most frequently prescribed combinations (as confirmed by practicing clinicians working with sepsis patients) and employ these for separate therapy options. These combinations are the following: all combinations of Linezolid or Vancomycin with Cefazolin, Cefepime, Cefotaxim, Cefpodoxim, Ceftazidim, Ceftriaxon, Cefuroxim, Piperacillin/Tazobactam, Ampicillin/Sulbactam, Oxacillin, Moxifloxacin, Levofloxacin, Ciprofloxacin, Imipenem, Meropenem, Ertapenem; all combinations of Clarithromycin with Ceftriaxon, Ceftazidim, Cefotaxim, Piperacillin/Tazobactam, Ampicillin/Sulbactam; all combinations of Tigecyclin with Levofloxacin and Ciprofloxacin; and the combination of Piperacillin/Tazobactam with Levofloxacin. We merge resistance results of combined therapies in such manner that the combined result is resistant if both single antibiotics are resistant, susceptible if at least one therapy is susceptible and missing if either both therapies have missing results or one is missing and one is resistant.

We remove the following therapies since they would not be administered in practice at the hospital which provided the data: Erythromycin, Tetracyclin, Aztreonam, Fusidinsaeure, Teicoplanin, Daptomycin. In addition, we only include therapies and combinations that have been tested in at least 200 patients in order to focus on the most common therapies and reduce the imbalance between classes. For the application of all HRS we remove all antibiotic therapies which have less than ten observations either with result ‘R’ or ‘S’ in AST. Finally, we exclude the combination therapies in the analysis since all of them have a high class imbalance between the resistance classes. All single and combined therapies are shown in Table A1.

### A.2 Data Preprocessing

#### A.2.1 Model-specific data

##### Imputation of missing values

For the model-based Approach IIb, which includes patient core variables in prediction models, data without missing values is required. Figure A1 shows that a complete case analysis is not feasible with the current dataset. Therefore we apply data imputation to the missing values. However, we remove observations which contain missing values for the suspected infection focus since this variable already contains a high level of uncertainty due to its derivation from diagnose codes. Therefore it seems too unreliable to impute values for this variable. This reduces the total sample size for the analysis with Approach IIb to 600 patients. For all remaining variables from core data, vital signs and blood values, we perform median imputation, which is, according to Choi et al. (2024), a simple but conservative approach comparable to more complex methods, and we consider it sufficient for our study as data imputation is not the focus of this work and solely required for Approach IIb. After imputation, we normalize all numeric variables from core data, vital signs and blood values to mean zero and standard deviation one.

**Figure A2:**
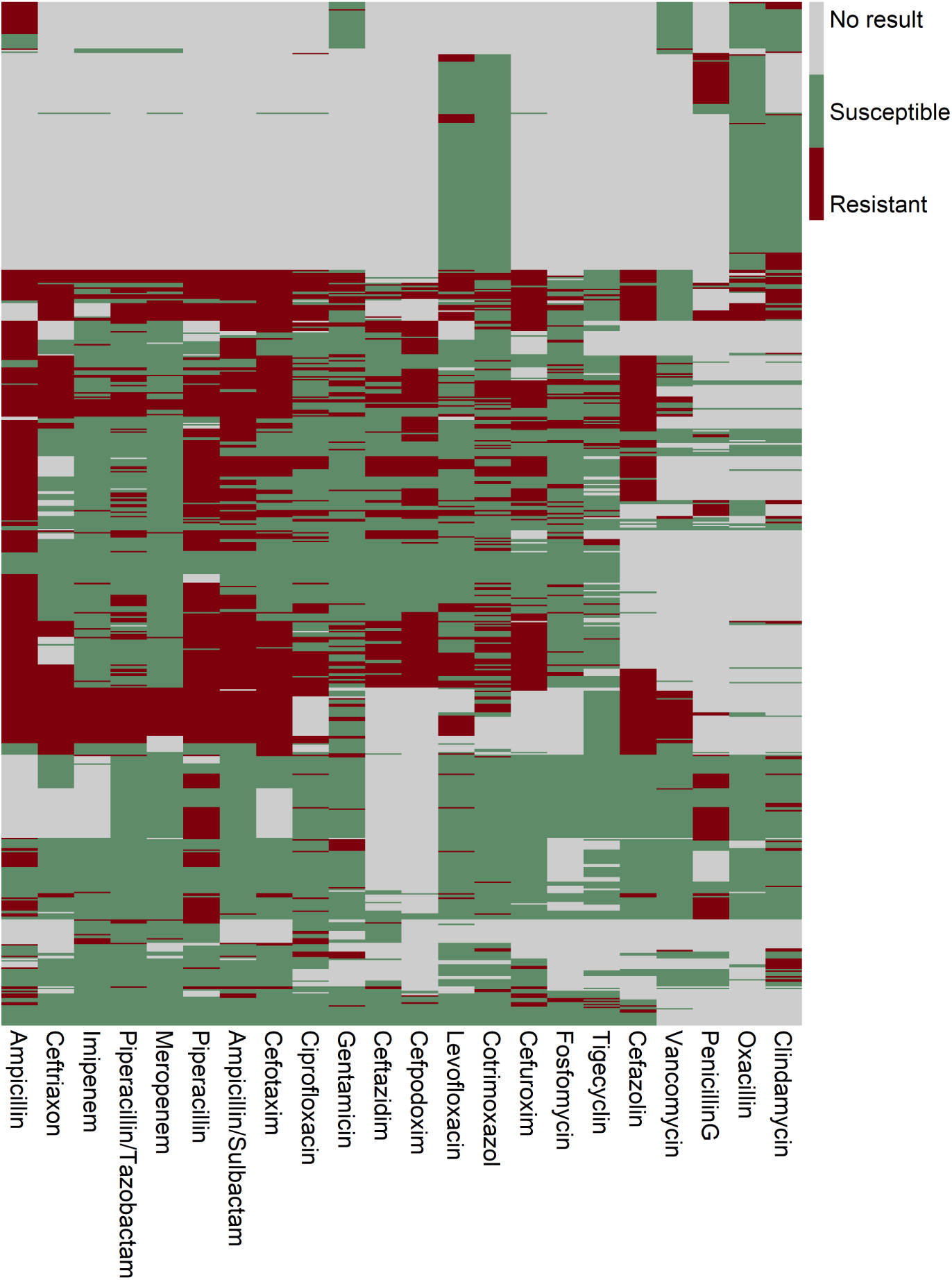
Graphical representation of AST results: observations (infection IDs) are shown in rows, therapies (single and combined) in columns. Only therapies that are included in the final analyses are shown. Red color marks resistant results, green color susceptible results and gray color indicates that no testing is done for the according combination of patient and therapy.

**Table A1:**
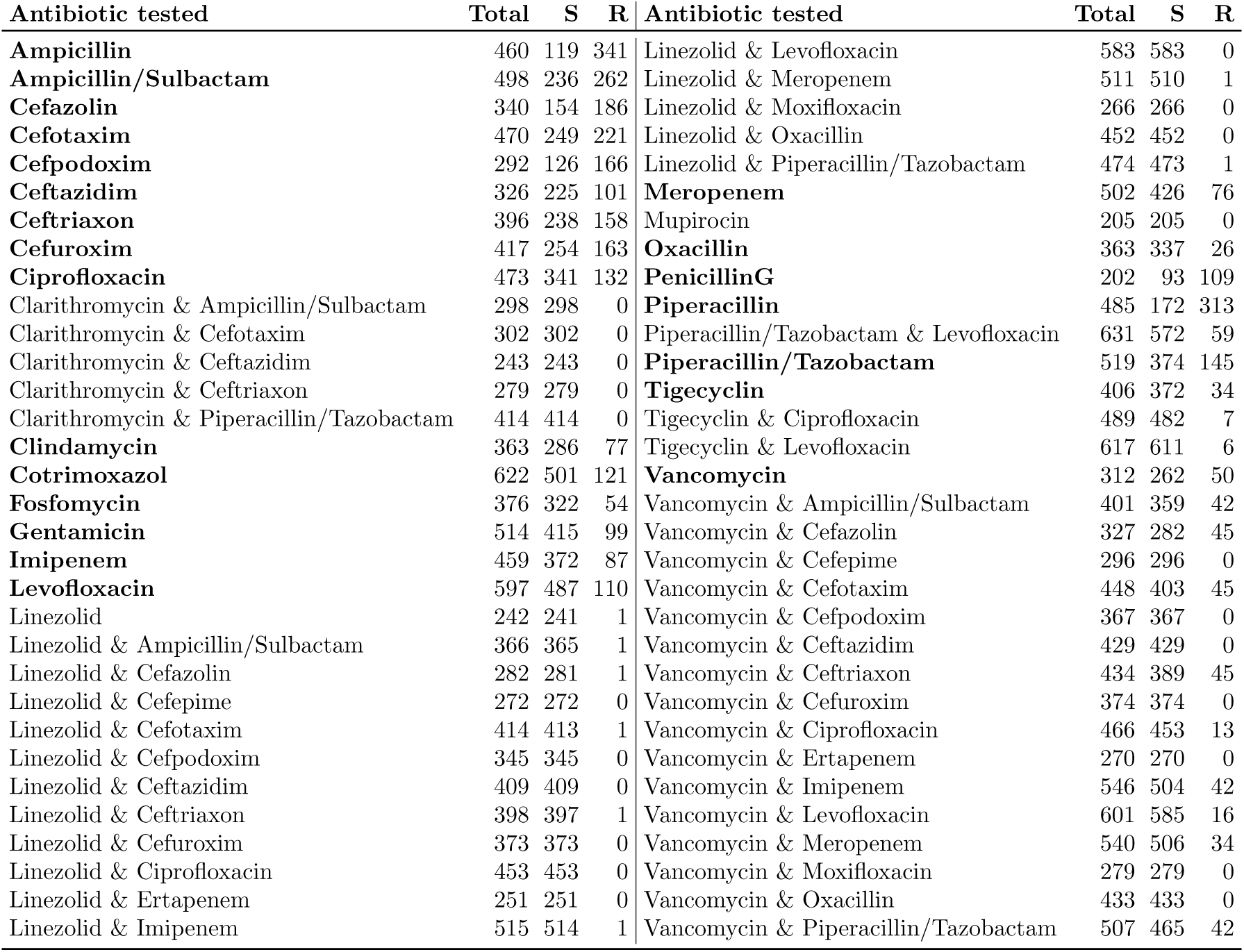
Frequencies of total prescriptions and susceptible (S) and resistant (R) AST results per tested antibiotic and relevant combinations over 703 patients. Antibiotics printed in bold are included in the final analysis.

| Antibiotic tested | Total | S | R | Antibiotic tested | Total | S | R |
| --- | --- | --- | --- | --- | --- | --- | --- |
| **Ampicillin** | 460 | 119 | 341 | Linezolid & Levofloxacin | 583 | 583 | 0 |
| **Ampicillin/Sulbactam** | 498 | 236 | 262 | Linezolid & Meropenem | 511 | 510 | 1 |
| **Cefazolin** | 340 | 154 | 186 | Linezolid & Moxifloxacin | 266 | 266 | 0 |
| **Cefotaxim** | 470 | 249 | 221 | Linezolid & Oxacillin | 452 | 452 | 0 |
| **Cefpodoxim** | 292 | 126 | 166 | Linezolid & Piperacillin/Tazobactam | 474 | 473 | 1 |
| **Ceftazidim** | 326 | 225 | 101 | **Meropenem** | 502 | 426 | 76 |
| **Ceftriaxon** | 396 | 238 | 158 | Mupirocin | 205 | 205 | 0 |
| **Cefuroxim** | 417 | 254 | 163 | **Oxacillin** | 363 | 337 | 26 |
| **Ciprofloxacin** | 473 | 341 | 132 | **PenicillinG** | 202 | 93 | 109 |
| Clarithromycin & Ampicillin/Sulbactam | 298 | 298 | 0 | **Piperacillin** | 485 | 172 | 313 |
| Clarithromycin & Cefotaxim | 302 | 302 | 0 | Piperacillin/Tazobactam & Levofloxacin | 631 | 572 | 59 |
| Clarithromycin & Ceftazidim | 243 | 243 | 0 | **Piperacillin/Tazobactam** | 519 | 374 | 145 |
| Clarithromycin & Ceftriaxon | 279 | 279 | 0 | **Tigecyclin** | 406 | 372 | 34 |
| Clarithromycin & Piperacillin/Tazobactam | 414 | 414 | 0 | Tigecyclin & Ciprofloxacin | 489 | 482 | 7 |
| **Clindamycin** | 363 | 286 | 77 | Tigecyclin & Levofloxacin | 617 | 611 | 6 |
| **Cotrimoxazol** | 622 | 501 | 121 | **Vancomycin** | 312 | 262 | 50 |
| **Fosfomycin** | 376 | 322 | 54 | Vancomycin & Ampicillin/Sulbactam | 401 | 359 | 42 |
| **Gentamicin** | 514 | 415 | 99 | Vancomycin & Cefazolin | 327 | 282 | 45 |
| **Imipenem** | 459 | 372 | 87 | Vancomycin & Cefepime | 296 | 296 | 0 |
| **Levofloxacin** | 597 | 487 | 110 | Vancomycin & Cefotaxim | 448 | 403 | 45 |
| Linezolid | 242 | 241 | 1 | Vancomycin & Cefpodoxim | 367 | 367 | 0 |
| Linezolid & Ampicillin/Sulbactam | 366 | 365 | 1 | Vancomycin & Ceftazidim | 429 | 429 | 0 |
| Linezolid & Cefazolin | 282 | 281 | 1 | Vancomycin & Ceftriaxon | 434 | 389 | 45 |
| Linezolid & Cefepime | 272 | 272 | 0 | Vancomycin & Cefuroxim | 374 | 374 | 0 |
| Linezolid & Cefotaxim | 414 | 413 | 1 | Vancomycin & Ciprofloxacin | 466 | 453 | 13 |
| Linezolid & Cefpodoxim | 345 | 345 | 0 | Vancomycin & Ertapenem | 270 | 270 | 0 |
| Linezolid & Ceftazidim | 409 | 409 | 0 | Vancomycin & Imipenem | 546 | 504 | 42 |
| Linezolid & Ceftriaxon | 398 | 397 | 1 | Vancomycin & Levofloxacin | 601 | 585 | 16 |
| Linezolid & Cefuroxim | 373 | 373 | 0 | Vancomycin & Meropenem | 540 | 506 | 34 |
| Linezolid & Ciprofloxacin | 453 | 453 | 0 | Vancomycin & Moxifloxacin | 279 | 279 | 0 |
| Linezolid & Ertapenem | 251 | 251 | 0 | Vancomycin & Oxacillin | 433 | 433 | 0 |
| Linezolid & Imipenem | 515 | 514 | 1 | Vancomycin & Piperacillin/Tazobactam | 507 | 465 | 42 |

##### Handling class imbalance in therapies

Table A1 shows that for many therapies there is a (very) high class imbalance between the two resistance classes ‘S’ and ‘R’. Ignoring this imbalance during modeling might lead to reduced classification performance (Huang and Dai, 2021). To account for this issue during the modeling stage of Approaches IIa and IIb, we use class weights in the fitting process of mboost. The idea is to assign larger weights to the minority class and lower weights to the majority class, so that the model pays more attention to the under-represented class during training. The weights are inversely proportional to the class frequencies and used in the function argument ‘weights’.

### A.3 Details on methods

#### Relationship of (C)DSS and (H)RS

DSS and RS are closely related. Both are designed to help users by filtering information, but with different approaches: DSS enable users to make their own decisions by providing analysis and information, whereas an RS actively guides users by recommending options based on anticipated preferences. As an example, in the context of searching for a new car, a DSS would provide information on various models, including comparisons of attributes such as cost, features, and fuel consumption, allowing the user to make an informed decision. In contrast, an RS would analyze the user’s preferences, past purchases, and the preferences of other (similar) users to proactively offer a personalized recommendation for a specific car. Such an RS could be incorporated as a tool within a DSS.

**Figure A3:**
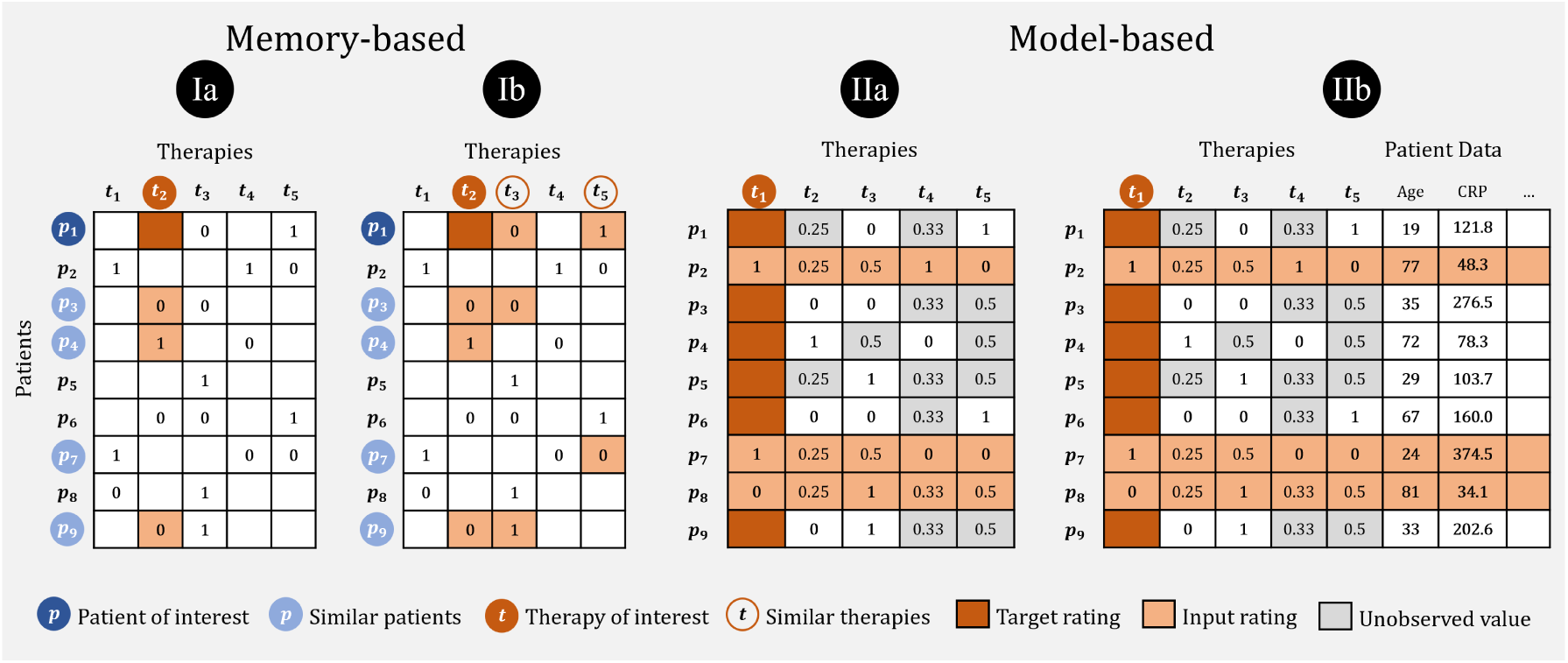
Graphical overview of all four different approaches for HRS. **Ia:** Hybrid memory-based user-based HRS; **Ib:** Hybrid memory-based user- and item-based HRS; **IIa:** Hybrid model-based HRS; **IIb:** Hybrid model-based HRS integrating patient covariates.

HRS are a subgroup of RS that give recommendations in the healthcare context. In that case, the role of the users is that of patients, and the items translate to diagnoses, treatments, health services, or personalized diets (Cai et al., 2022). Clinical decision support systems CDSS are DSS in the clinical context. HRS can be incorporated into clinical workflows within a CDSS to offer personalized recommendations, for example regarding therapies, thereby contributing to the overall decision support objectives.

Figure A3 provides an overview about the four developed HRS approaches.

#### Similarities and rating matrix

In order to calculate the similarity between patients by using Gower’s distance (see Equation (1)), weights *w_v_* are set for the patient variables. The weights result from medical knowledge and experience of the clinical work with patients suffering from sepsis. The importance of each variable is expressed on a scale between 1 (less important) and 5 (most important), in terms of its relevance in defining similar patients. The weights for the variables, which are also listed in table 1, are as follows: Age (3), Sex (1), Suspected infection focus (5), GCS (3), Heart rate (3), mIbp (3), Bilirubin (3), fio2 (5), pO2 (4), Leucocytes (4), Thrombocytes (3), CRP (4), GFR (4), Creatinine (4), IL6 (4), Lactate (5), Blood sugar (2), Body temperature (4). Following, all pairwise similarities between patients are calculated.

To calculate similarity between therapies, we first need information about pathogen coverage for all therapies. Since we use data from a German hospital, we employ the AMBOSS antibiotic mosaic (AMBOSS GmbH, 2024) as basis. Coverage of antibiotics which are not listed in the mosaic were additionally discussed with medical experts. Finally, a pathogen-therapy matrix is created including the expected effectiveness of all antibiotics included in the data against specific pathogens.

Additionally, we build the patient-therapy matrix (rating matrix), including all patients and their AST results. The matrix contains values of 0 (resistant) and 1 (susceptible) as a result for therapies that were tested with AST for the respective patient, and missing values otherwise (see Figure A2).

### A.4 Details on results

The following Figures A4 to A7 and Tables A2 to A5 provide additional information to the results presented in Section 3.

**Figure A4:**
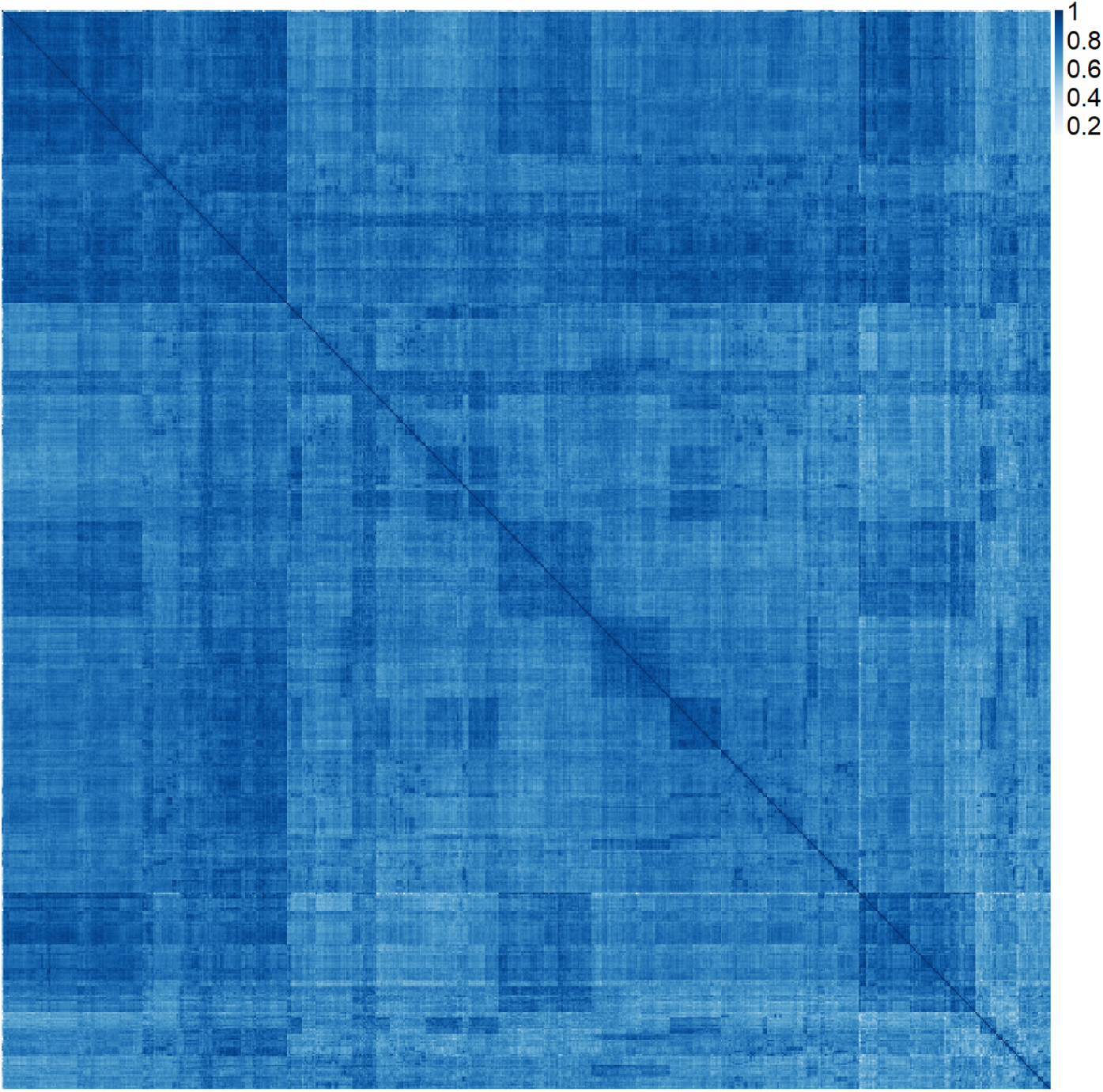
Heatmap of pairwise patient similarities calculated according to Equation (1). The horizontal and vertical axis contain all patients in the same order. The color key goes from white (low similarity) to dark blue (high similarity). The values off the main diagonal (which display a patient’s similarity with themselves) range from 0.12 to 0.996 (mean 0.76, median 0.76). High similarity may, among others, be caused by the fact that some patients appear multiple times in the data for multiple independent infections.

**Figure A5:**
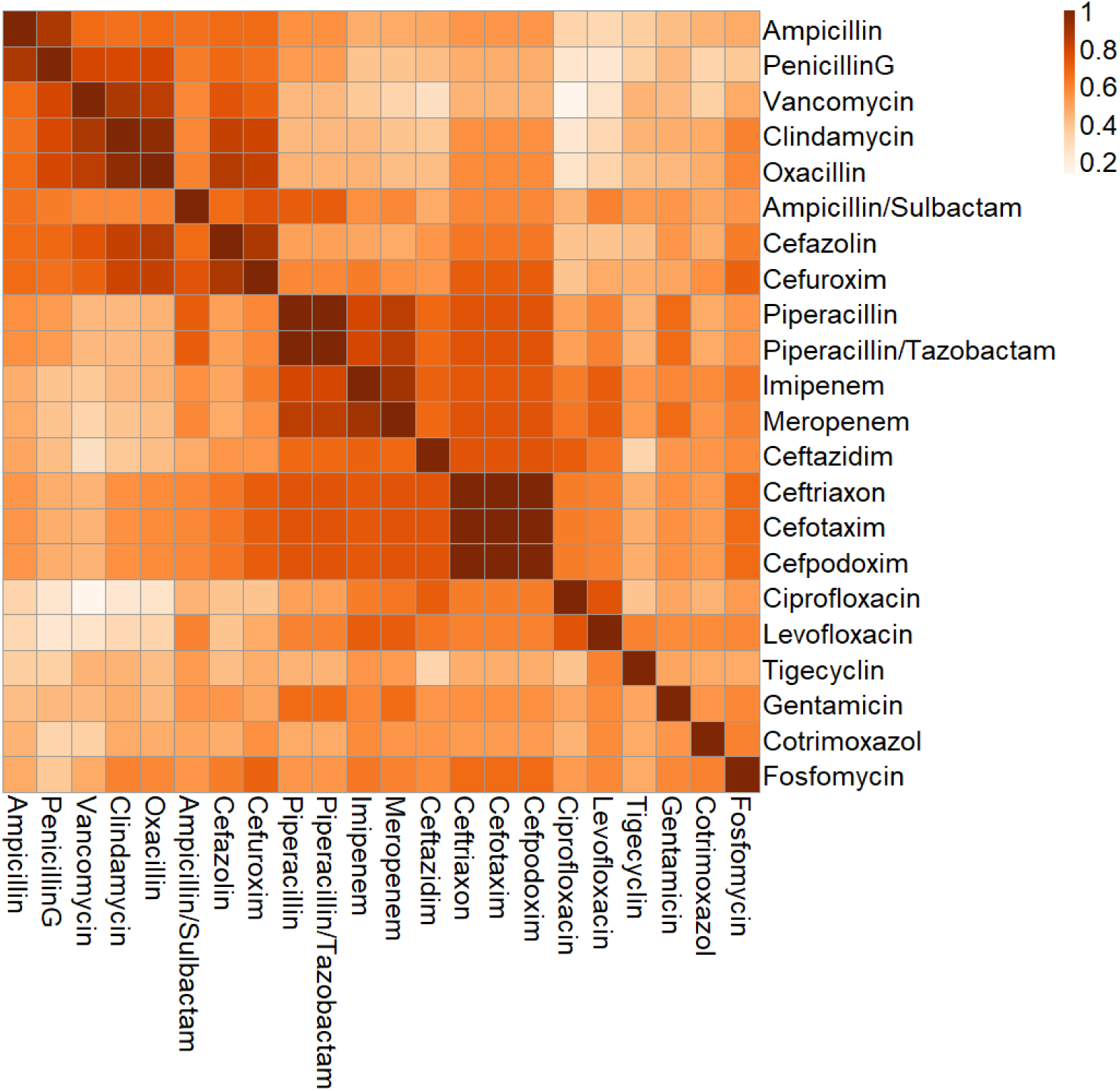
Heatmap of pairwise therapy similarities calculated according to Equation (5). The color key goes from white (low similarity) to dark red (high similarity). Apparently, there are groups of therapies within which therapies are more similar to each other than to other therapies. The displayed values (excluding the self-similarities on the main diagonal) range from 0.13 to 1.00 (mean 0.57, median 0.56).

**Table A2:** Results of Approach Ia for various choices of the number *ν* of similar patients or the patient similarity threshold *ρ*, each for both splitting strategies. Performance measures AUC, sensitivity, specificity, precision and F1 score are shown. The results with the highest F1 score for each combination are highlighted in bold. For equal values of F1 score, precision became relevant.

| Splitting | Patient similarity | AUC | Sensitivity | Specificity | Precision | F1 |
| --- | --- | --- | --- | --- | --- | --- |
| Row-wise | $\nu = 10$ | 0.690 | 0.790 | 0.487 | 0.763 | 0.775 |
| | $\nu = 50$ | 0.736 | 0.867 | 0.457 | 0.769 | 0.814 |
| | $\nu = 100$ | **0.742** | **0.887** | **0.432** | **0.765** | **0.820** |
| | $\nu = 250$ | 0.742 | 0.897 | 0.395 | 0.756 | 0.819 |
| | $\nu = 500$ | 0.742 | 0.878 | 0.419 | 0.759 | 0.813 |
| | $\nu = 1000$ | 0.741 | 0.860 | 0.453 | 0.766 | 0.810 |
| | $\rho = 0.95$ | 0.628 | 0.646 | 0.560 | 0.725 | 0.681 |
| | $\rho = 0.90$ | 0.673 | 0.807 | 0.459 | 0.759 | 0.781 |
| | $\rho = 0.85$ | 0.729 | 0.875 | 0.426 | 0.762 | 0.814 |
| | $\rho = 0.80$ | **0.744** | **0.894** | **0.413** | **0.761** | **0.821** |
| | $\rho = 0.75$ | 0.742 | 0.881 | 0.416 | 0.759 | 0.814 |
| | $\rho = 0.70$ | 0.741 | 0.864 | 0.441 | 0.763 | 0.809 |
| | $\rho = 0.65$ | 0.741 | 0.861 | 0.451 | 0.766 | 0.810 |
| | $\rho = 0.60$ | 0.741 | 0.860 | 0.452 | 0.766 | 0.809 |
| Cell-wise | $\nu = 10$ | 0.686 | 0.784 | 0.483 | 0.762 | 0.772 |
| | $\nu = 50$ | 0.736 | 0.867 | 0.453 | 0.769 | 0.815 |
| | $\nu = 100$ | **0.741** | **0.883** | **0.428** | **0.764** | **0.819** |
| | $\nu = 250$ | 0.740 | 0.895 | 0.392 | 0.755 | 0.819 |
| | $\nu = 500$ | 0.739 | 0.878 | 0.416 | 0.759 | 0.814 |
| | $\nu = 1000$ | 0.738 | 0.859 | 0.454 | 0.768 | 0.810 |
| | $\rho = 0.95$ | 0.615 | 0.649 | 0.554 | 0.723 | 0.683 |
| | $\rho = 0.90$ | 0.667 | 0.805 | 0.448 | 0.755 | 0.779 |
| | $\rho = 0.85$ | 0.727 | 0.872 | 0.426 | 0.763 | 0.813 |
| | $\rho = 0.80$ | **0.742** | **0.894** | **0.411** | **0.761** | **0.822** |
| | $\rho = 0.75$ | 0.739 | 0.880 | 0.412 | 0.758 | 0.815 |
| | $\rho = 0.70$ | 0.738 | 0.863 | 0.442 | 0.764 | 0.810 |
| | $\rho = 0.65$ | 0.738 | 0.860 | 0.452 | 0.767 | 0.811 |
| | $\rho = 0.60$ | 0.738 | 0.859 | 0.453 | 0.767 | 0.810 |

**Table A3:** Results of Approach Ib for various choices of the therapy similarity threshold *τ* and for both splitting strategies. Patient similarity thresholds were set to *ν* = 100 and *ρ* = 0.8, respectively, which arose as optimal choices in Approach Ia. Performance measures AUC, sensitivity, specificity, precision and F1 score are shown. The results with the highest precision and F1 score for each combination are highlighted in bold.

| Splitting | Patient sim. | Therapy sim. | AUC | Sensitivity | Specificity | Precision | F1 |
| --- | --- | --- | --- | --- | --- | --- | --- |
| Row-wise | $\nu = 100$ | $\tau = 1$ | **0.733** | **0.908** | **0.343** | **0.742** | **0.816** |
| | | $\tau = 0.95$ | **0.731** | **0.908** | **0.343** | **0.742** | **0.816** |
| | | $\tau = 0.9$ | **0.730** | **0.908** | **0.343** | **0.742** | **0.816** |
| | | $\tau = 0.85$ | 0.719 | 0.907 | 0.329 | 0.738 | 0.813 |
| | | $\tau = 0.8$ | 0.707 | 0.925 | 0.277 | 0.727 | 0.813 |
| | | $\tau = 0.75$ | 0.696 | 0.939 | 0.240 | 0.721 | 0.815 |
| | | $\tau = 0.7$ | 0.694 | 0.971 | 0.160 | 0.707 | 0.817 |
| | $\rho = 0.80$ | $\tau = 1$ | **0.735** | **0.928** | **0.303** | **0.736** | **0.820** |
| | | $\tau = 0.95$ | **0.733** | **0.928** | **0.303** | **0.736** | **0.820** |
| | | $\tau = 0.9$ | **0.732** | **0.928** | **0.303** | **0.736** | **0.820** |
| | | $\tau = 0.85$ | 0.721 | 0.929 | 0.288 | 0.731 | 0.817 |
| | | $\tau = 0.8$ | 0.711 | 0.937 | 0.258 | 0.725 | 0.816 |
| | | $\tau = 0.75$ | 0.702 | 0.951 | 0.221 | 0.718 | 0.818 |
| | | $\tau = 0.7$ | 0.699 | 0.975 | 0.145 | 0.704 | 0.817 |
| Cell-wise | $\nu = 100$ | $\tau = 1$ | **0.731** | **0.905** | **0.335** | **0.741** | **0.814** |
| | | $\tau = 0.95$ | **0.729** | **0.905** | **0.335** | **0.741** | **0.814** |
| | | $\tau = 0.9$ | **0.729** | **0.905** | **0.335** | **0.741** | **0.814** |
| | | $\tau = 0.85$ | 0.718 | 0.904 | 0.323 | 0.737 | 0.812 |
| | | $\tau = 0.8$ | 0.707 | 0.923 | 0.276 | 0.728 | 0.814 |
| | | $\tau = 0.75$ | 0.696 | 0.937 | 0.239 | 0.721 | 0.815 |
| | | $\tau = 0.7$ | 0.693 | 0.970 | 0.157 | 0.707 | 0.818 |
| | $\rho = 0.80$ | $\tau = 1$ | **0.735** | **0.930** | **0.304** | **0.737** | **0.822** |
| | | $\tau = 0.95$ | **0.732** | **0.930** | **0.304** | **0.737** | **0.822** |
| | | $\tau = 0.9$ | **0.733** | **0.930** | **0.304** | **0.737** | **0.822** |
| | | $\tau = 0.85$ | 0.722 | 0.929 | 0.292 | 0.734 | 0.820 |
| | | $\tau = 0.8$ | 0.713 | 0.938 | 0.258 | 0.726 | 0.818 |
| | | $\tau = 0.75$ | 0.705 | 0.952 | 0.221 | 0.720 | 0.820 |
| | | $\tau = 0.7$ | 0.704 | 0.975 | 0.146 | 0.706 | 0.819 |

**Table A4:** Results of Approaches IIa and IIb. Performance measures AUC, sensitivity, specificity, precision and F1 score are shown for different combinations of splitting strategy, basis for initialization and matrix update. When Approach Ia is used as basis for initial values, the patient similarity threshold is set to *ρ* = 0.8. For Approach Ib as basis, it is set to *ρ* = 0.8 and the therapy similarity threshold to *τ* = 0.90.

| Splitting | HRS | Basis | Matrix update | AUC | Sensitivity | Specificity | Precision | F1 |
| --- | --- | --- | --- | --- | --- | --- | --- | --- |
| Row-wise | IIa | Ia | collective | 0.576 | 0.486 | 0.580 | 0.708 | 0.574 |
|  | IIa | Ia | immediate | 0.582 | 0.334 | 0.712 | 0.706 | 0.448 |
|  | IIa | Ib | collective | 0.576 | 0.232 | 0.779 | 0.702 | 0.321 |
|  | IIa | Ib | immediate | 0.589 | 0.459 | 0.618 | 0.716 | 0.555 |
|  | IIb | Ia | collective | 0.547 | 0.488 | 0.474 | 0.657 | 0.558 |
|  | IIb | Ia | immediate | 0.539 | 0.468 | 0.506 | 0.662 | 0.544 |
|  | IIb | Ib | collective | 0.535 | 0.415 | 0.528 | 0.643 | 0.502 |
|  | IIb | Ib | immediate | 0.541 | 0.482 | 0.485 | 0.660 | 0.553 |
| Cell-wise | IIa | Ia | collective | 0.903 | 0.799 | 0.859 | 0.922 | 0.856 |
|  | IIa | Ia | immediate | 0.904 | 0.795 | 0.858 | 0.922 | 0.853 |
|  | IIa | Ib | collective | 0.904 | 0.793 | 0.859 | 0.922 | 0.852 |
|  | IIa | Ib | immediate | 0.902 | 0.795 | 0.856 | 0.921 | 0.853 |
|  | IIb | Ia | collective | 0.902 | 0.817 | 0.850 | 0.918 | 0.864 |
|  | IIb | Ia | immediate | 0.904 | 0.811 | 0.854 | 0.920 | 0.862 |
|  | IIb | Ib | collective | 0.900 | 0.810 | 0.847 | 0.916 | 0.859 |
|  | IIb | Ib | immediate | 0.902 | 0.813 | 0.854 | 0.920 | 0.863 |

**Table A5:** Performance results as in Table 4, this time for cell-wise splitting.

| Therapy | Ia |  |  | Ib |  |  | IIa |  |  | IIb |  |  |
| --- | --- | --- | --- | --- | --- | --- | --- | --- | --- | --- | --- | --- |
|  | Sens | Spec | F1 | Sens | Spec | F1 | Sens | Spec | F1 | Sens | Spec | F1 |
| Ampicillin | 0.00 | 1.00 | - | 0.00 | 1.00 | - | 0.87 | 0.89 | 0.80 | 0.85 | 0.87 | 0.76 |
| **AmpSul** | **0.64** | **0.57** | **0.59** | **0.64** | **0.57** | **0.59** | **0.91** | **0.90** | **0.90** | **0.92** | **0.90** | **0.90** |
| **Cefazolin** | **0.58** | **0.69** | **0.59** | **0.58** | **0.69** | **0.59** | **0.97** | **0.91** | **0.93** | **0.97** | **0.90** | **0.91** |
| **Cefotaxim** | **0.81** | **0.35** | **0.67** | **0.86** | **0.33** | **0.69** | **0.98** | **0.95** | **0.97** | **0.99** | **0.95** | **0.97** |
| **Cefpodoxim** | **0.12** | **0.92** | **0.22** | **0.83** | **0.30** | **0.59** | **0.90** | **0.83** | **0.85** | **0.89** | **0.86** | **0.85** |
| Ceftazidim | 0.98 | 0.01 | 0.81 | 0.98 | 0.01 | 0.81 | 0.79 | 0.95 | 0.87 | 0.78 | 0.87 | 0.85 |
| **Ceftriaxon** | **0.95** | **0.16** | **0.75** | **0.87** | **0.36** | **0.75** | **0.95** | **1.00** | **0.97** | **0.97** | **0.98** | **0.98** |
| Cefuroxim | 0.96 | 0.12 | 0.76 | 0.96 | 0.12 | 0.76 | 0.94 | 0.92 | 0.94 | 0.90 | 0.94 | 0.92 |
| Ciprofloxacin | 1.00 | 0.00 | 0.84 | 1.00 | 0.00 | 0.84 | 0.86 | 0.78 | 0.89 | 0.88 | 0.82 | 0.90 |
| Clindamycin | 1.00 | 0.00 | 0.88 | 1.00 | 0.00 | 0.88 | 0.56 | 0.64 | 0.66 | 0.77 | 0.53 | 0.81 |
| Cotrimoxazol | 1.00 | 0.00 | 0.89 | 1.00 | 0.00 | 0.89 | 0.67 | 0.77 | 0.77 | 0.66 | 0.79 | 0.77 |
| Fosfomycin | 1.00 | 0.00 | 0.92 | 1.00 | 0.00 | 0.92 | 0.60 | 0.61 | 0.72 | 0.62 | 0.74 | 0.74 |
| Gentamicin | 1.00 | 0.00 | 0.89 | 1.00 | 0.00 | 0.89 | 0.67 | 0.69 | 0.77 | 0.66 | 0.63 | 0.76 |
| Imipenem | 1.00 | 0.00 | 0.90 | 1.00 | 0.00 | 0.90 | 0.88 | 0.87 | 0.92 | 0.92 | 0.85 | 0.94 |
| Levofloxacin | 1.00 | 0.00 | 0.90 | 1.00 | 0.00 | 0.90 | 0.74 | 0.89 | 0.83 | 0.80 | 0.85 | 0.87 |
| Meropenem | 1.00 | 0.00 | 0.92 | 1.00 | 0.00 | 0.92 | 0.91 | 0.98 | 0.95 | 0.94 | 0.94 | 0.96 |
| Oxacillin | 1.00 | 0.00 | 0.96 | 1.00 | 0.00 | 0.96 | 0.75 | 0.93 | 0.85 | 0.81 | 0.92 | 0.89 |
| **PenicillinG** | **0.54** | **0.45** | **0.49** | **0.54** | **0.45** | **0.49** | **0.85** | **0.71** | **0.77** | **0.84** | **0.67** | **0.73** |
| Piperacillin | 0.00 | 0.99 | - | 0.98 | 0.12 | 0.53 | 0.90 | 0.82 | 0.81 | 0.92 | 0.87 | 0.84 |
| PipTaz | 1.00 | 0.00 | 0.84 | 0.94 | 0.14 | 0.82 | 0.84 | 0.83 | 0.88 | 0.80 | 0.86 | 0.86 |
| Tigecyclin | 1.00 | 0.00 | 0.96 | 1.00 | 0.00 | 0.96 | 0.65 | 0.74 | 0.77 | 0.65 | 0.49 | 0.76 |
| Vancomycin | 1.00 | 0.00 | 0.91 | 1.00 | 0.00 | 0.91 | 0.78 | 0.84 | 0.85 | 0.75 | 0.76 | 0.83 |

**Figure A6:**
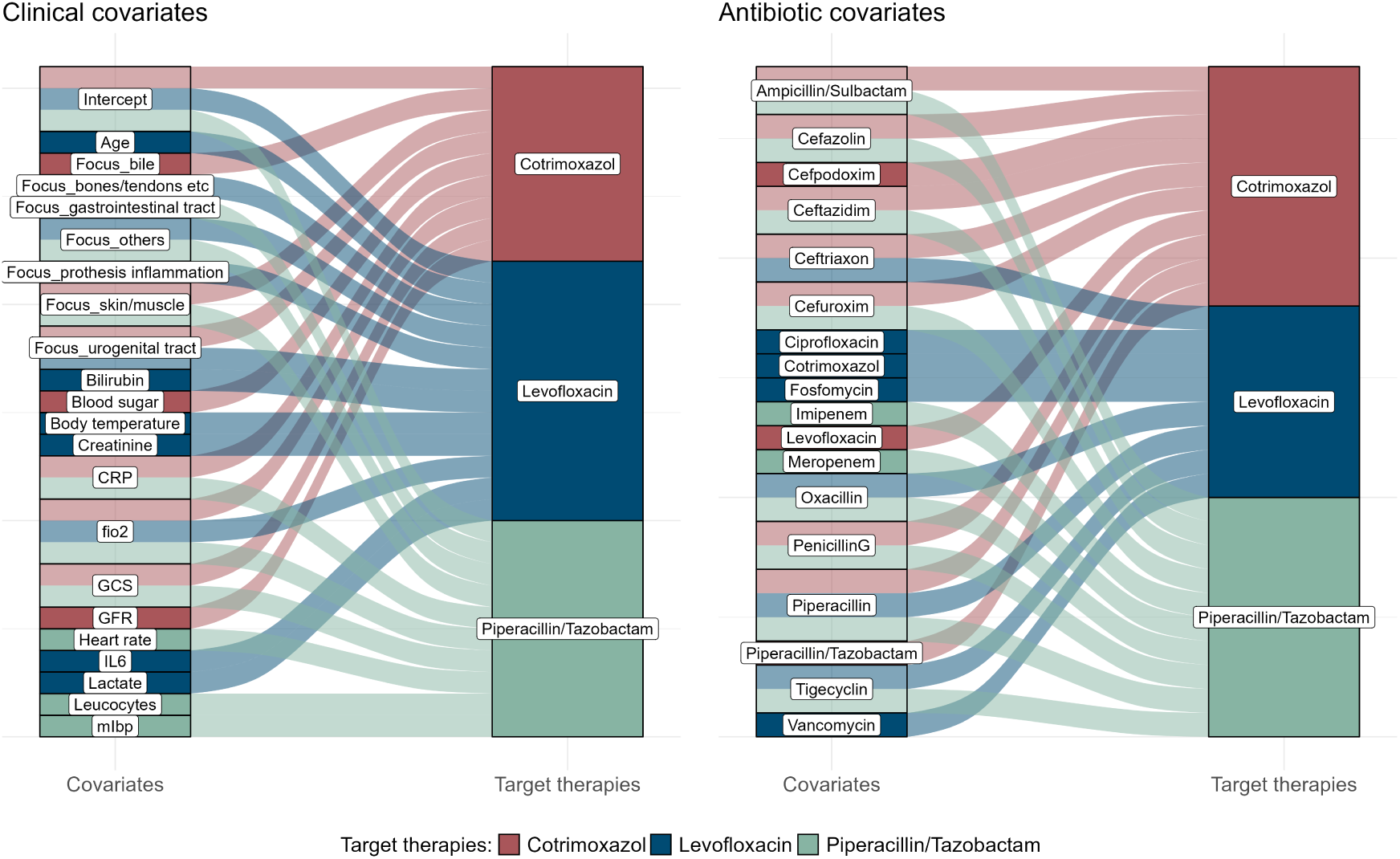
Covariates chosen by model: Exemplary illustration of variables which are identified by the mboost algorithm as covariates for prediction of the therapy response of specific antibiotic therapies. The HRS in the displayed example uses Approach IIb with patient-therapy matrix values initialized through Approach Ib with immediate matrix update and the mboost model for classification. To maintain clarity, only the three most frequently tested single therapies (Cotrimoxazol, Levofloxacin, Piperazillin/Tazobactam) are shown here. Each variable is linked to those therapies for which it is chosen as a covariate to predict the therapy response. On the left side, all clinical variables (from core data, vital signs and blood values) are listed, while the antibiotic covariates are shown on the right. The purpose of this distinction is to provide a more comprehensive overview.

**Figure A7:**
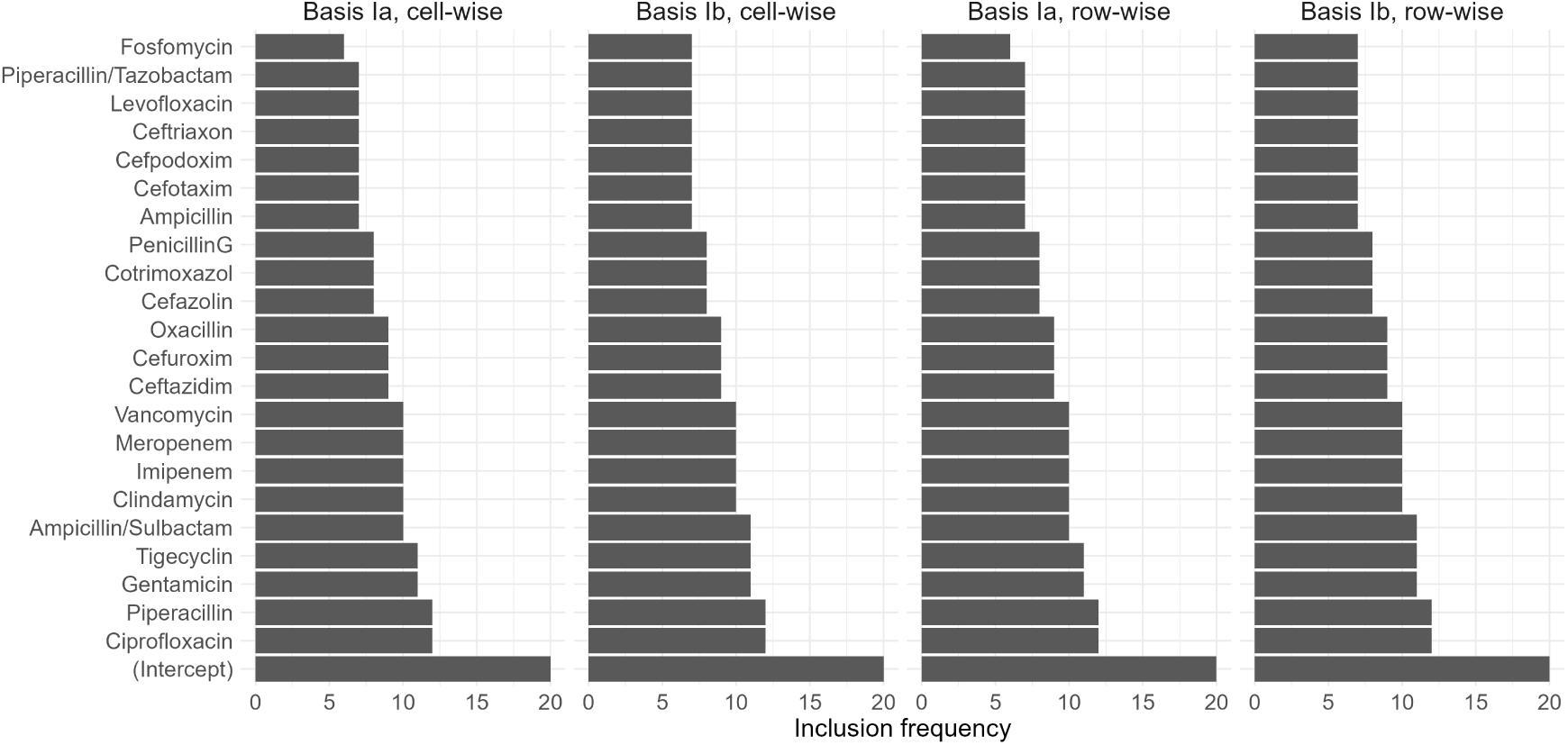
Selection frequency (over all target therapies) of covariates within the mboost models of Approach IIa with immediate matrix update, both for the models which use Approach Ia and Ib as basis for initialization, and for both cell-wise and row-wise splitting.

